# Trans-ancestry meta-analysis improves performance of genetic scores for multiple adiposity-related traits in East Asian populations

**DOI:** 10.1101/2022.07.05.22277254

**Authors:** Zammy Fairhurst-Hunter, Kuang Lin, Iona Y Millwood, Alfred Pozarickij, Tzu-Ting Chen, Jason M. Torres, Jian-an Lun, Christiana Kartsonaki, Wei Gan, Anubha Mahajan, Huaidong Du, Rajani Sohoni, Yu Guo, Sam Sansome, Ling Yang, Canqing Yu, Yiping Chen, Jun Lv, Gibran Hemani, Masaru Koido, Yoichiro Kamatani, Cassandra N. Spracklen, Penny Gordon-Larsen, Mine Koprulu, Xiangrui Meng, Karoline Kuchenbaecker, Segun Fatumo, Laxmi Bhatta, Ben Brumpton, Jesús Alegre-Díaz, Pablo Kuri-Morales, Roberto Tapia-Conyer, Sarah E. Graham, Cristen J. Willer, Matt J. Neville, Fredrik Karpe, Mariaelisa Graff, Kari E North, Ruth J.F. Loos, Christopher A. Haiman, Ulrike Peters, Steve Buskye, Christopher R. Gignoux, Genevieve Wojcik, Yen-Fen Ling, Liming Li, Mark I McCarthy, Zhengming Chen, Michael V Holmes, Robin G Walters

**Affiliations:** Clinical Trial Service Unit and Epidemiological Studies Unit (CTSU), Nuffield Department of Population Health, University of Oxford, Oxford, UK; Medical Research Council Population Health Research Unit (MRC PHRU), Nuffield Department of Population Health, University of Oxford, Oxford, UK; Center for Neuropsychiatric Research, National Health Research Institutes, Miaoli, Taiwan; MRC Epidemiology Unit, Institute of Metabolic Science, University of Cambridge School of Clinical Medicine, Cambridge, UK; Wellcome Centre for Human Genetics, Nuffield Department of Medicine, University of Oxford, Oxford, UK; Chinese Academy of Medical Sciences, Beijing, China; Department of Epidemiology and Biostatistics, School of Public Health, Peking University, Beijing, China; MRC Integrative Epidemiology Unit, University of Bristol, Bristol, UK; Graduate School of Frontier Sciences, The University of Tokyo, Minato-ku, Japan; Biostatistics and Epidemiology, University of Massachusetts Amherst, Amherst; Nutrition Department, Gillings School of Global Public Health, UNC-Chapel Hill, Chapel Hill; Division of Psychiatry, University College London, London, UK; The African Computational Genomics (TACG) Research Group, MRC/UVRI and LSHTM, Entebbe, Uganda; The Department of Non-communicable Disease Epidemiology, London School of Hygiene and Tropical Medicine, London, UK; K.G. Jebsen Center for Genetic Epidemiology, Department of Public Health and Nursing, Norwegian University of Science and Technology NTNU, Trondheim, Norway; Experimental Research Unit from the Faculty of Medicine (UIME), National Autonomous University of Mexico (UNAM), Mexico City, Mexico; Department of Internal Medicine, Division of Cardiology, University of Michigan, Ann Arbor, US; Oxford NIHR Biomedical Research Centre, Oxford University Hospitals Trust, Oxford, UK; Oxford Centre for Diabetes, Endocrinology and Metabolism, Radcliffe Department of Medicine, University of Oxford, Oxford, UK; Department of Epidemiology, University of North Carolina, Chapel Hill, North Carolina; The Charles Bronfman Institute of Personalized Medicine, Icahn School of Medicine at Mount Sinai, Mount Sinai, New York City, US; Center for Genetic Epidemiology, Keck School of Medicine, University of Southern California, Los Angeles, US; Division of Public Health Science, Fred Hutchinson Cancer Research Center, Seattle, US; Department of Statistics, Rudgers University, New Brunswick; Colorado Center for Personalized Medicine, University of Colorado Anschutz Medical Campus, Aurora; Division of Genetic Epidemiology, John Hopkins University, Baltimore; Department of Public Health & Medical Humanities, School of Medicine, National Yang Ming Chiao Tung University, Taipei, Taiwan; Institute of Behavioral Medicine, College of Medicine, National Cheng Kung University, Tainan, Taiwan; Medical Research Council, Integrative Epidemiology Unit, University of Bristol, UK

## Abstract

Genome-wide association studies (GWAS) in predominately European-ancestry (EUR) populations have identified numerous genetic variants associated with adiposity-related traits. An emerging challenge is the limited transferability of genetic scores constructed based on GWAS results from one ancestry for trait prediction in other ancestries. We performed trans-ancestry meta-analysis (TAMA) for eight adiposity-related traits using genetic data from 96,124 East Asian (EAS) and 443,359 EUR individuals. We identified >1400 genomic regions significantly associated with one or more traits. Despite EAS comprising only ∼20% of the study population, genetic scores constructed from the trans-ancestry (TA) results accounted for between 30% and 79% more variation in the adiposity traits in EAS compared with scores derived from the EUR GWAS alone. Furthermore, TA scores also modestly improved variance explained in African/African American, Hispanic and South Asian populations. Our findings highlight the utility of TAMA for increasing variance explained by genetic scores across populations of different ancestries.

## Introduction

Overweight and obesity affects around 2 billion adults worldwide^1^, and its prevalence continues to rise steadily in most countries including China, where there has been an increase of ∼25% over the past two decades.^2^ Although the association between excess adiposity and risk of many diseases is well established, uncertainty remains about the underlying pathological mechanisms leading to disease. In particular, the relative utilities of different measures of adiposity (e.g. reflecting overall body fat versus its distribution) in predicting disease risk and their roles in disease pathophysiology are yet to be reliably established. Furthermore, there is emerging evidence that the relationship between adiposity measures and disease risk varies substantially by ancestry. For instance, individuals of East Asian (EAS) ancestry appear more susceptible to metabolic disorders at a lower body mass index (BMI) than is typically observed in European (EUR) ancestry populations^3, 4^, due perhaps to a greater propensity of EAS individuals to store fat centrally^5, 6^.

Genetic scores that are predictive of adiposity traits can be used as instrumental variables (IV) in Mendelian randomisation (MR) analysis for an unbiased assessment of the causal effects of adiposity measures on health^7, 8^. Typically, genetic scores have been constructed from genetic variants associated with the exposure of interest at genome-wide significance^9^, but, more recently, Bayesian and clumping/thresholding methods have been developed to derive polygenic scores (PGS) from genome-wide summary statistics that are much more predictive^10–12^. However, because these PGS include information from thousands to millions of variants that are mostly not significantly associated with the trait of interest, the variants comprising these scores are not appropriate for use in MR due to violation of the IV assumption of a robust association with the exposure of interest^8, 13, 14^. Furthermore, the use of PGS in MR can exacerbate the influence of weak instrument bias and winner’s curse^15, 16^ and methods for assessing horizontal pleiotropy and performing two-sample or multivariable MR are not easily applicable to PGS^17, 18^.

Large-scale genome-wide association studies (GWAS) in predominately EUR populations have assessed various anthropometric measures of adiposity^19–22^, providing a source of variants that can be combined into a score and utilised as an IV in MR. However, it is becoming increasingly apparent that genetic scores derived in one ancestry are less predictive and hence provide reduced statistical power for analyses of the corresponding trait or disease in populations of other ancestries^23–26^. Furthermore, given the over-representation of EUR in GWAS to date, there is concern that the poor transferability of scores between ancestries will not only lead to biased risk estimates but may also exacerbate existing health inequalities between populations^23, 27^. Notably, as the emergence of overweight/obesity in East Asia is a relatively recent phenomenon^2^, and the health effects have not been so extensively studied as in EUR, there is a clear need for genetic scores which are strong instruments of adiposity in EAS to facilitate detailed investigations of the impact of overweight/obesity in these populations. Although GWAS of adiposity traits have been conducted in EAS populations, well-powered studies have been reported for only a small number of adiposity traits, predominately focusing on overall adiposity measures rather than body fat distribution^28, 29^.

The present study seeks to characterise the genetic architecture of adiposity-related traits in EAS and EUR populations and to develop genetic scores that will facilitate future well-powered genetic studies investigating the health impact and relative importance in EAS, as well as in EUR, of overall adiposity and of body fat distribution. To achieve this, we performed GWAS and trans-ancestry meta-analysis (TAMA) for eight adiposity-related traits in the China Kadoorie Biobank (CKB) and the UK Biobank (UKB). We compared the heritability of each trait in the two populations and assessed the genetic overlap between adiposity traits. Finally, we constructed genetic scores derived from the UKB, CKB and trans-ancestry (TA) results and assessed their performance in a range of populations from multiple different ancestry groups.

## Results

### Distribution of anthropometric traits in UKB and CKB

Based on previously published work^30–32^, we selected eight traits related to different aspects of weight and body shape: BMI, body fat percentage (Fat%), waist circumference (WC), and hip circumference (HIP) were classed as measures of overall adiposity, while waist-to-hip ratio (WHR) and the derived parameters WC adjusted for BMI (WCadjBMI), HIP adjusted for BMI (HIPadjBMI), and WHR adjusted for BMI (WHRadjBMI) were considered to be more reflective of body fat distribution.

Mean values for BMI, Fat%, WC and HIP were appreciably higher in UKB than in CKB participants for both men and women (**Supplementary Table 1**). The mean value for WHR in UKB men was also higher than in CKB men (UKB 0.94 versus CKB 0.90) but the reverse was true for women (UKB 0.82 versus CKB 0.87). In both studies, mean values of BMI and HIP were similar between the sexes, and mean values for Fat% were 0.7-fold smaller in men then in women. In UKB, mean values of both WC and WHR were 15% larger in men than in women but in CKB the differences were only 3%.

In both populations the overall adiposity traits were highly correlated with one another (0.7≤*r*≤0.87) (**Supplementary Figure 1a and Supplementary Table 2**), the body fat distribution traits WHR, WHRadjBMI and WCadjBMI were also highly positively correlated with one another (0.61≤*r*≤0.85) but had a weaker correlation with HIPadjBMI (−0.38≤*r*≤0.31). Conversely, correlation between the overall adiposity traits and the body fat distribution traits in UKB and CKB were much weaker (−0.01≤*r*≤0.059), with the exception of the relationship of WC with WHR (*r*_UKB_ = 0.78, *r*_CKB_=0.83). Although the general patterns of cross-trait correlation were similar between the populations, for many of the comparisons there was a significant difference in *r* (P-values < 10^-6^). For instance, WHR had a stronger correlation with the overall adiposity traits and a weaker relationship with the other body fat distribution traits in CKB than in UKB. Similar patterns were observed when comparing the Pearson correlation values estimated separately by sex (**Supplementary Figure 1b**).

### GWAS of adiposity traits in UKB and CKB

GWAS of each of the eight traits were conducted in each biobank, using rank-inverse-normal-transformed traits. To take account of differences between sexes in the distributions of some of these traits (**Supplementary Table 1**), GWAS were run sex-stratified and results were meta-analysed using the Han and Eskin random effects (HE-RE) model to improve power to detect associations in the presence of effect size heterogeneity^33, 34^. All further downstream analyses of the GWAS results were restricted for each biobank to variants with minor allele frequency (MAF) ≥0.01 in that biobank and a genome-wide significance threshold of P<5×10^-8^ was applied.

Defining a locus as the region of the genome spanned by variants in linkage disequilibrium (LD) with a lead variant, with overlapping regions merged into a single larger locus, we identified between 400 to 675 loci significantly associated with each trait in UKB and between 21 to 71 loci in CKB (**Table 1**). Per trait, these loci included 480 to 789 and 22 to 73 conditionally-independent variants associated at P<5×10^-8^ in UKB and CKB, respectively (**Table 1, Supplementary Figures 2 and 3, Supplementary Data 1 and 2**). If instead the MAF threshold was lowered to 0.001 in UKB and 0.005 in CKB (the larger threshold in CKB reflecting the smaller sample size), and a correspondingly more stringent significance threshold of P<5×10^-9^ was applied, ∼25% and ∼33% fewer loci and conditionally-independent variants overall were identified in UKB and CKB, respectively (**Supplementary Table 3**). In CKB, no variants with MAF<0.01 were significantly associated with any of the traits, and in UKB only 2.3% or fewer of variants significantly associated with each trait had a MAF<0.01.

**Table 1).** Number of loci and conditionally independent variants significantly associated with the adiposity traits in the UKB, CKB or TA analyses

| Trait | UKB<br>N = 443,359 N variants = 9,737,881 |  |  | CKB<br>N = 96,124 N variants = 8,561,153 |  |  | TA<br>N = 539,483 N variants = 12,225,321 |  |  |
| --- | --- | --- | --- | --- | --- | --- | --- | --- | --- |
|  | Loci | Novel Loci | Conditionally independent variants | Loci | Novel Loci | Conditionally independent variants | Loci | Novel Loci | Conditionally independent variants |
| BMI | 675 | 21 | 789 | 71 | 6 | 73 | 816 | 52 | 907 |
| Fat% | 617 | 533 | 712 | 40 | 22 | 40 | 692 | 599 | 768 |
| WC | 510 | 458 | 575 | 46 | 29 | 47 | 604 | 548 | 633 |
| HIP | 619 | 545 | 740 | 45 | 25 | 45 | 709 | 629 | 804 |
| WHR | 405 | 53 | 480 | 22 | 5 | 22 | 439 | 78 | 504 |
| WCadjBMI | 512 | 79 | 674 | 35 | 5 | 35 | 567 | 99 | 681 |
| HIPadjBMI | 663 | 555 | 927 | 61 | 28 | 61 | 757 | 637 | 960 |
| WHRadjBMI | 400 | 37 | 528 | 21 | 3 | 22 | 459 | 57 | 537 |

### Trans-ancestry meta-analysis

For each trait, we conducted TAMA using the summary statistics from the biobank-specific, sex-stratified GWAS (i.e. combining four separate GWAS per trait), again applying the HE-RE model to allow for both ancestry- and sex-heterogeneity in variant effect size. Due to varying patterns of LD in different ancestries, current methods for locus definition based on LD and for identification of conditionally-independent signals are not readily applicable to results from TAMA. To address this, we developed a pipeline to incorporate local LD structure from both EUR and EAS (**see Supplementary Methods, Supplementary Figures 4 and 5**). Using this pipeline, between 439 to 816 loci and 504 to 960 conditionally independent variants were significantly (P<5×10^-8^) associated with each trait in the trans-ancestry (TA) results (**Table 1, Supplementary Figure 6, Supplementary Data Table 3**).

After harmonising locus boundaries from the UKB, CKB, and TA analyses by combining sets of overlapping loci into single merged loci for each trait separately, TAMA was found to identify an additional 56 to 111 loci per trait over those found in the biobank-specific analyses (**Figure 1a**). A small proportion (0.1% to 11% per trait) of loci associated in each individual biobank did not reach genome wide significance in the TAMA (**Figure 1a**); on inspection these were mainly associations that only marginally surpassed genome-wide significance in one biobank while lying closely around the null in the other (**Supplementary Table 4, Supplementary Figures 7a and 8a**). We checked the effect sizes for the lead variants at these UKB- and CKB-specific loci in previous studies from the GIANT consortium^22, 35^ and Taiwan Biobank (TWB)^28^, respectively. Effect sizes of variants identified at the UKB-specific loci were consistently smaller when estimated in GIANT than in UKB, likely due to the influence of winner’s curse (**Supplementary Figure 7b**). However, across the traits, few variants per trait (<6%) showed signs of heterogeneity and the majority of effects (>92%) were in the same direction. Conversely for the same variants in CKB, no more than 56% had effect sizes in the same direction as in UKB and up to 65% showed signs of heterogeneity (**Supplementary Figure 7a**). We observed a similar pattern of results for the CKB-specific loci comparisons; effect sizes were more similar between CKB and TWB than between CKB and UKB (**Supplementary Figure 8b**).

**Figure 1).**
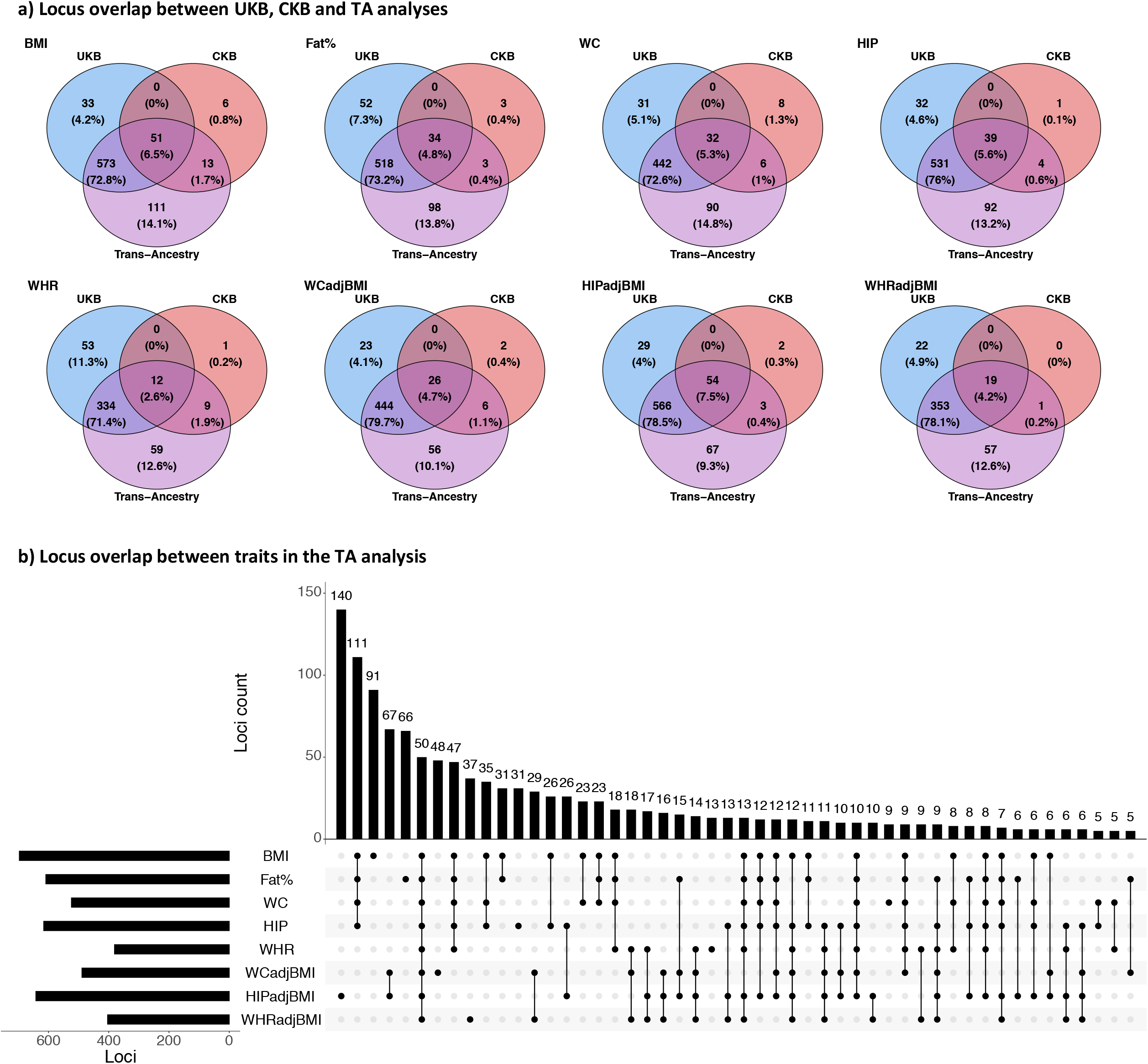
Positional overlap of loci between analyses and traits We compared the bp overlap of loci either between the UKB, CKB and TA results (1a) or between loci associated with any of the eight traits in the TAMA (1b). The central bar chart in 1B shows the number of genomic regions that were associated with a given combination of traits. These results are plotted only for the first 50 combinations of traits, ordered by number of associated genomic regions. The total number of loci associated with each trait is shown in the bar chart on the left.

For each analysis (UKB/CKB/TA) of each trait, we identified as novel those loci within which no previous genome-wide significant association signal had been reported by the GIANT consortium or otherwise documented in the GWAS catalog^20–22, 35–38^ (**Table 1**). For well-studied traits such as BMI, only a small proportion of loci were found to be novel but, for other traits that have not been studied in detail using GWAS results with large sample sizes, the majority of loci identified have not been previously reported. For instance, nearly 90% of the 709 loci associated with HIP were novel.

### Multi-trait associations

To assess the extent to which genomic regions were associated with multiple traits, locus boundaries from the eight TA analyses were harmonised by merging all overlapping loci, for all traits, into larger ‘regions of association’. There were 1,441 distinct regions of the genome showing association with one or more of the adiposity traits (**Figure 1b**), of which only 50 were associated with all eight traits. We found that 469 regions were associated with one or more of the overall adiposity traits but not with any of the body fat distribution traits, of which 111 were shared across all four overall adiposity traits. Conversely, of 420 regions associated with one or more of the fat distribution traits but with none of the overall adiposity traits, just 14 were shared by all four traits.

Using FUMA^39^, we performed gene-set enrichment and tissue expression analyses based on combined sets of conditionally-independent variants within the regions of the genome associated with either the overall adiposity traits or the body fat distribution traits. Genes located within 10Kbp up- or downstream of index variants in overall adiposity regions showed significant differential expression only in brain tissue (**Supplementary Figure 9a),** but genes at body fat distribution regions showed differential expression in a wide range of tissues, including in subcutaneous and visceral omentum adipose tissue (**Supplementary Figure 9b**). Overall adiposity genes showed enrichment for gene-sets relating to the brain and nervous system, with 43% of significant gene-sets containing terms relating to neuronal processes (**Supplementary Table 5**). Conversely, only 3% of the gene-sets that the body fat distribution genes were enriched for contained terms relating to neuronal processes and, instead, 55% of the gene-sets contained terms relating to development and morphology (**Supplementary Table 5**). When tissue specificity was assessed for each adipose trait separately, we observed the same pattern: genes lying in close proximity to index signals for the overall adiposity traits showed upregulated expression predominately in brain tissue, but body fat distribution traits showed a much broader tissue expression profile (**Supplementary Figures 10-11**).

### Heritability and genetic correlation

We estimated SNP-based heritability (h^2^_SNP_) of the eight traits using LD score regression (LDSC)^40^. Heritability was similar between the biobanks for traits classed as reflecting overall adiposity (h^2^_SNP_ 0.19-0.23 in UKB and in 0.17-0.22 CKB) but, for all the body fat distribution traits, heritability was significantly (P-values < 10^-5^) lower in CKB (h^2^_SNP_ 0.09-0.13) than in UKB (h^2^_SNP_ 0.14-0.20) (**Figure 2a and Supplementary Table 6**). This difference was even greater when comparing the female-specific h^2^_SNP_ estimates (h^2^_SNP_ 0.12-0.14 in CKB and 0.19-0.22 in UKB; P-values < 10^-4^) (**Supplementary Figure 12a**).

**Figure 2).**
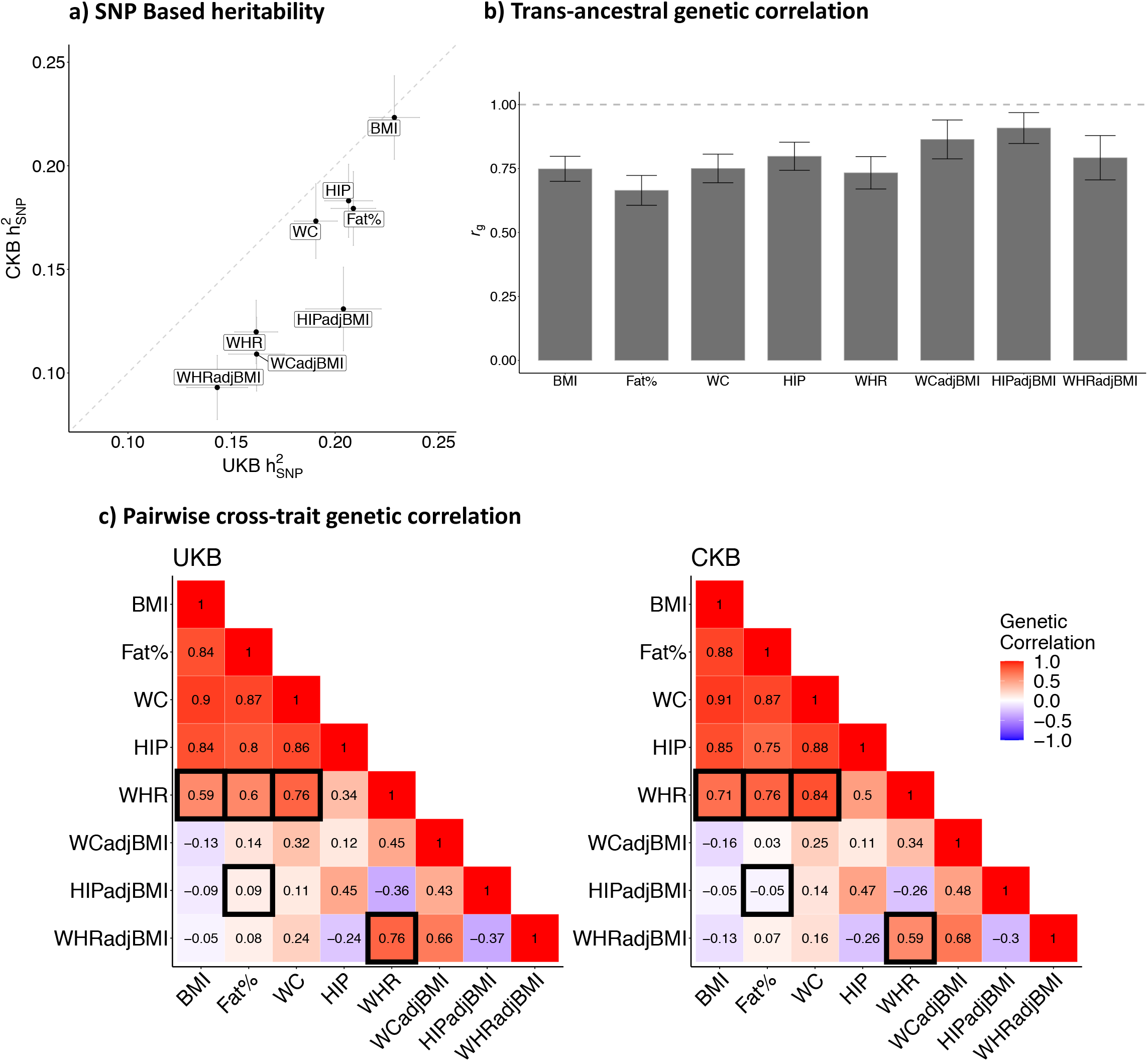
Assessment of the genetic architecture of the adiposity traits in UKB and CKB 2a) SNP based heritability estimated from the sex-combined summary statistics using LDSC regression, error bars represent the 95% confidence intervals (CI). 2b) Trans-ancestral genetic correlation between UKB and CKB estimated using Popcorn, error bars represent the 95% CI. 2c) Cross-trait genetic correlation estimated in UKB and CKB using LDSC regression, black boxes indicate a significant difference (All P-values < 10^-3^) in *r* between CKB and UKB. The alpha threshold was Bonferroni corrected by the 28 cross-trait comparisons made.

Sex-specific h^2^_SNP_ values estimated in UKB were largely the same between men and women for all traits other than WHR and WHRadjBMI, which showed greater heritability in women (**Supplementary Figure 12a**). However, after accounting for multiple testing this difference was only significant for WHRadjBMI (UKB Women h^2^_snp_ 0.20, [SE 0.013]; UKB Men h^2^_SNP_ 0.13, [SE 0.006]; P=6.1×10^-7^). Conversely in CKB there was no significant difference in h^2^_SNP_ values between the sexes for any of the traits.

Despite some differences in heritability, the traits showed consistently high cross-ancestry genetic correlations (*r*_g_) between UKB and CKB as measured using Popcorn^41^ (*r*_g_ 0.66-0.91, median 0.77) (**Figure 2b, Supplementary Table 7**). Nevertheless, *r*_g_ was significantly lower than 1 for all traits (P-values < 0.003). We observed similar results when assessing *r*_g_ using the sex-specific GWAS results (**Supplementary Figure 12b**).

We also examined pairwise genetic correlations between the eight traits in each biobank (**Figure 2c, Supplementary Table 8**). The overall adiposity traits showed strong positive genetic correlation with one another in both studies (*r*_g_ > 0.75). WHR also showed strong genetic correlation with the overall adiposity traits, but this relationship was stronger in CKB than in UKB, particularly for BMI, Fat% and WC (P-values < 10^-3^). Conversely, cross-trait genetic correlation between WHR and WHRadjBMI was significantly higher in UKB (*r*_g_=0.76) than in CKB (*r*_g_=0.59) (P=2.1×10^-7^). These differences between the biobanks remained significant when comparing the cross-trait genetic correlations in UKB and CKB women, but were not different for the comparison of *r*_g_ values between UKB and CKB men (**Supplementary Figure 12c**).

### Performance of genetic scores

We constructed genetic scores for each of the traits based on the conditionally independent variants identified from the UKB, CKB, or TA analyses and tested their performance in UKB and CKB (**Figure 3 and Supplementary Figure 13**). In UKB, the UKB and TA scores performed similarly for each trait, accounting for 5%-12% of trait variance; CKB scores performed very poorly, however, with partial *r*^2^ values of only 0.3-1.0 (**Figure 3 and Supplementary Figure 13**). By contrast, in CKB the TA scores out-performed both CKB and UKB scores, accounting for 2%-6% of trait variance, while the CKB and UKB scores performed similarly, each accounting for 1%-4%. In both UKB and CKB, the most trait variation was accounted for by scores for HIPadjBMI and the smallest by those for WHR. When the scores were tested for each sex separately in the two biobanks, the trait variance accounted for by the scores was similar between the sexes, with the exception of WHR and WHRadjBMI; for these partial *r*^2^ values estimated in women were substantially larger than those estimated in men (2.3-2.9-fold greater for UKB, 2.1-2.6-fold for CKB) (**Supplementary Figure 14**).

**Figure 3).**
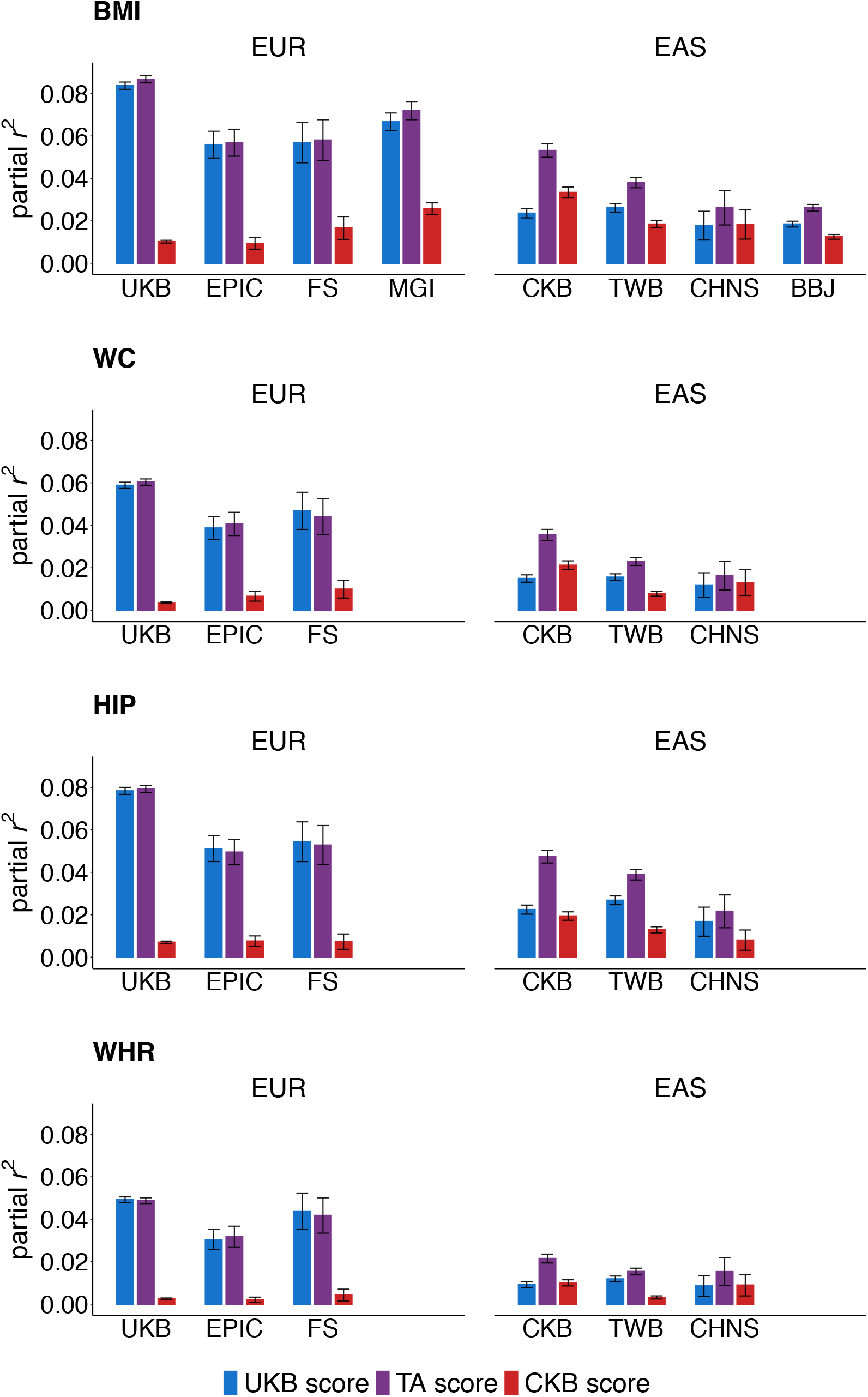
Score performance in EUR and EAS populations Partial *r*^2^ values for BMI, WC, HIP and WHR were estimated in the unrelated subsets of EUR or EAS populations from the regression of each trait against the corresponding scores with adjustment for study specific PCs and covariates, age, age^2^ and sex. The 95% CI are plotted based on the standard error of the partial *r*^2^ values estimated using Olkin and Finn’s approximation.

We additionally tested the scores for a subset of the traits (BMI, WC, HIP, and WHR) in multiple independent populations of EUR and EAS ancestry (**Figure 3**), in which the patterns of variance explained across the different score types were similar to those observed in UKB and CKB. However, for each trait the maximum proportion of variation explained by the scores was somewhat lower than in UKB and CKB; up to 7% and 4% of trait variance was accounted for in these independent EUR and EAS populations respectively.

While the TA and UKB scores performed very similarly in the EUR populations, with the TA score at most accounting for 1.09-fold more variation in the adiposity traits than the UKB scores, in EAS the improvement in the performance of the TA scores relative to the UKB scores was much more pronounced. Despite only a modest (9% to 22%) increase in the total number of variants included in the TA scores, partial *r*^2^ values in the independent EAS populations were 1.3- to 1.8-fold greater than those estimated using the UKB scores (**Figure 3**). Nevertheless, although there was a large improvement in variance explained for EAS when using the TA scores, these partial *r*^2^ values were still 0.4- to 0.7-fold lower than those estimated in EUR.

To better understand the poor transferability of genetic scores between ancestries we examined in more detail some of the contributing factors, using the UKB scores, which accounted for 0.3- to 0.5-fold less trait variance in EAS than in EUR, as an example. First, the variance explained by a genetic score is limited by the MAF of its constituent variants, which can vary considerably between populations^42^. Power for GWAS discovery is greater for more common variants, hence those variants identified in UKB GWAS exhibited a skew towards higher MAF in EUR, while there is no such skew for the same variants in EAS (**Supplementary Figure 15a**). Ancestry differences in patterns of LD can also impact whether variants detected in GWAS correctly tag the underlying causal variants, potentially reducing the utility of the tag variants for TA trait prediction. To illustrate, at the *MC4R* locus the patterns of LD vary considerably between EUR and EAS, with many more variants in high LD with the causal variant in EUR than in EAS (**Supplementary Figure 15b**). Using a method that quantifies the contribution of MAF and LD differences to variation in score performance between ancestries^42^, we predicted these factors combined would reduce the proportion of variation accounted for by the UKB scores in EAS by 0.7-fold compared with in EUR (**Figure 4a**). As such, MAF and LD may account for up to 59% of the reduced performance of the UKB scores in EAS compared with that in EUR (**Supplementary Table 9**), with the remaining reduction in performance most likely attributable to differences in causal effect sizes and heritability.

**Figure 4).**
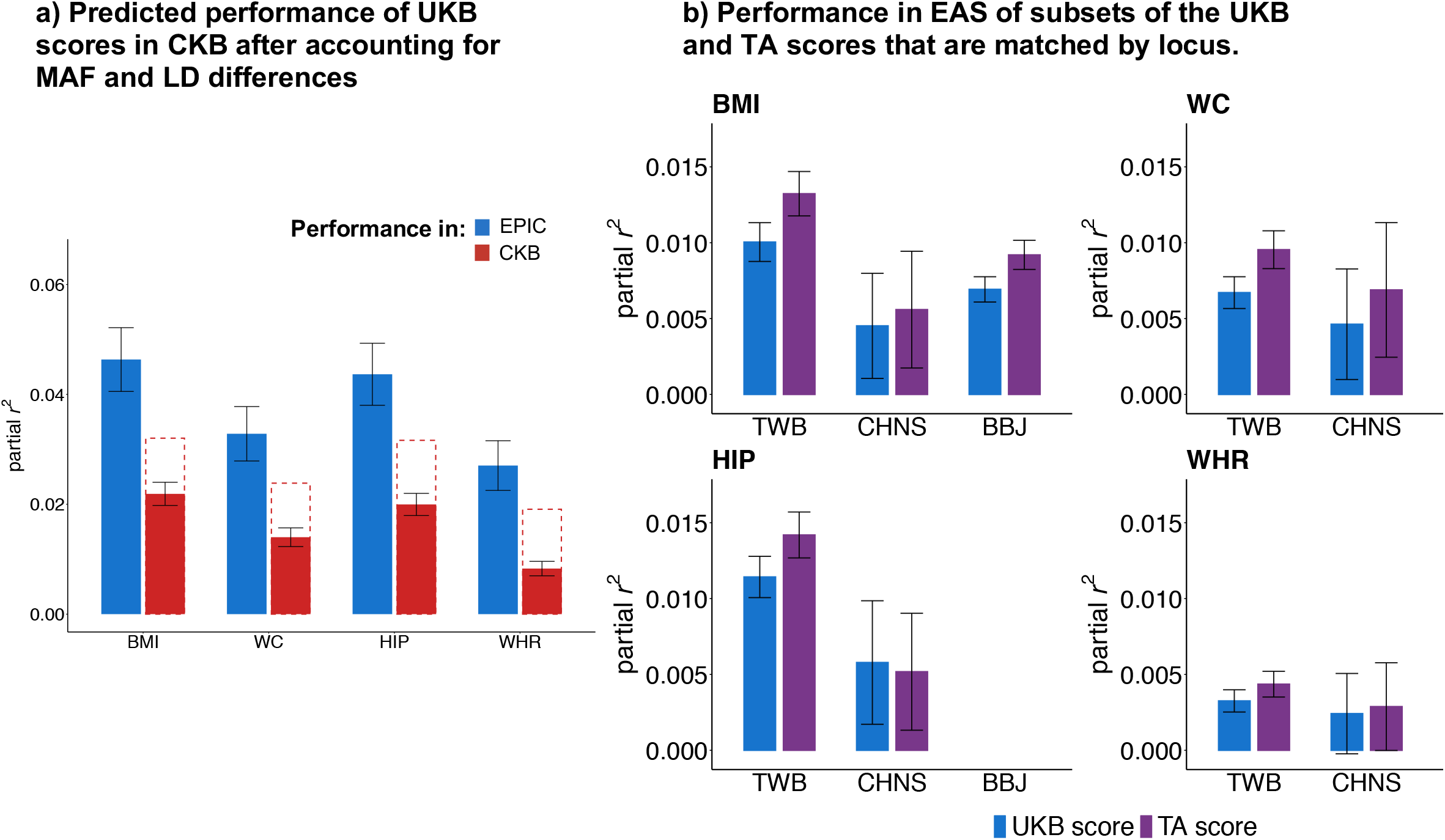
Factors affecting the transferability of genetic scores between ancestries 4a) Comparison of the performance of UKB scores in EPIC (EUR) and CKB (EAS). Based on the performance of the scores in EPIC, we estimated the partial *r*^2^ values that would be expected in CKB after accounting for MAF and LD differences between EAS and EUR, which we plot as the dashed red line. 4b) To directly compare how the TAMA influences the selection of variants, we created subsets of the UKB and TA scores that represented the same regions of the genome and contained the same number of variants. We weighted the scores using effect sizes estimated in UKB and tested them in independent EAS cohorts. The 95% CI are plotted based on the standard error of the partial *r*^2^ values estimated using Olkin and Finn’s approximation.

Given the predicted impact of LD on the transferability of genetic scores, where causal variants are shared between ancestries but local LD structure differs, TAMA may have the potential to identify variants that act as better tags in both populations, thereby improving score transferability. Accordingly, at some loci we observed that TAMA resulted in a change in lead variant from that selected initially in the UKB analysis (**Supplementary Figure 16**). To evaluate whether this change represented the selection of better tagging variants, we constructed scores using only a subset of loci for which the variants identified from the conditional analysis differed between the UKB and TA results. Despite the scores for each trait including the same number of variants, the partial *r*^2^ values estimated in EAS populations for the TA subset scores were on average 1.27-fold larger than those estimated for the UKB subset scores (**Figure 4b**). By contrast, in the EUR replication cohorts there was no difference in variance explained between the subsetted UKB and TA scores (**Supplementary Figure 17**).

### Genetic score performance in other ancestries

We additionally assessed the relative performance of UKB, CKB and TA scores for BMI, WC, HIP and WHR in populations of African/African American (AFR/AA), South Asian (SAS), or Hispanic (HIS) ancestry (**Figure 5**). In all populations and for almost all traits, the variance explained by the TA scores was consistently larger than for the UKB scores, although the observed proportional increase in *r*^2^ varied by ancestry. For instance, across the traits the variance explained by TA scores were on average 1.16-fold greater than for the UKB scores in individuals with AFR/AA ancestry, but only 1.06-fold greater in SAS. In HIS participants from The Population Architecture through Genomics and Environment study (PAGE)^43^, partial *r*^2^ values estimated for the TA scores were on average 1.09-fold greater than those estimated for the UKB scores, while in HIS participants of the Mexico City Prospective Study (MCPS)^44^, the mean improvement in variance explained was 1.18-fold. The difference in improvement between the two HIS populations may reflect differences in population characteristics and environmental exposure but it may also relate to the lower mean proportion of estimated EUR ancestry (28.2%) for MCPS participants than for HIS PAGE participants (54%). For a subset of MCPS participants with a particularly low proportion of EUR ancestry (EUR ancestry <20%, N=12,662), the improvement in variance explained by the TA scores compared to UKB scores was even greater (1.12- to 1.34-fold) (**Supplementary Figure 18**).

**Figure 5).**
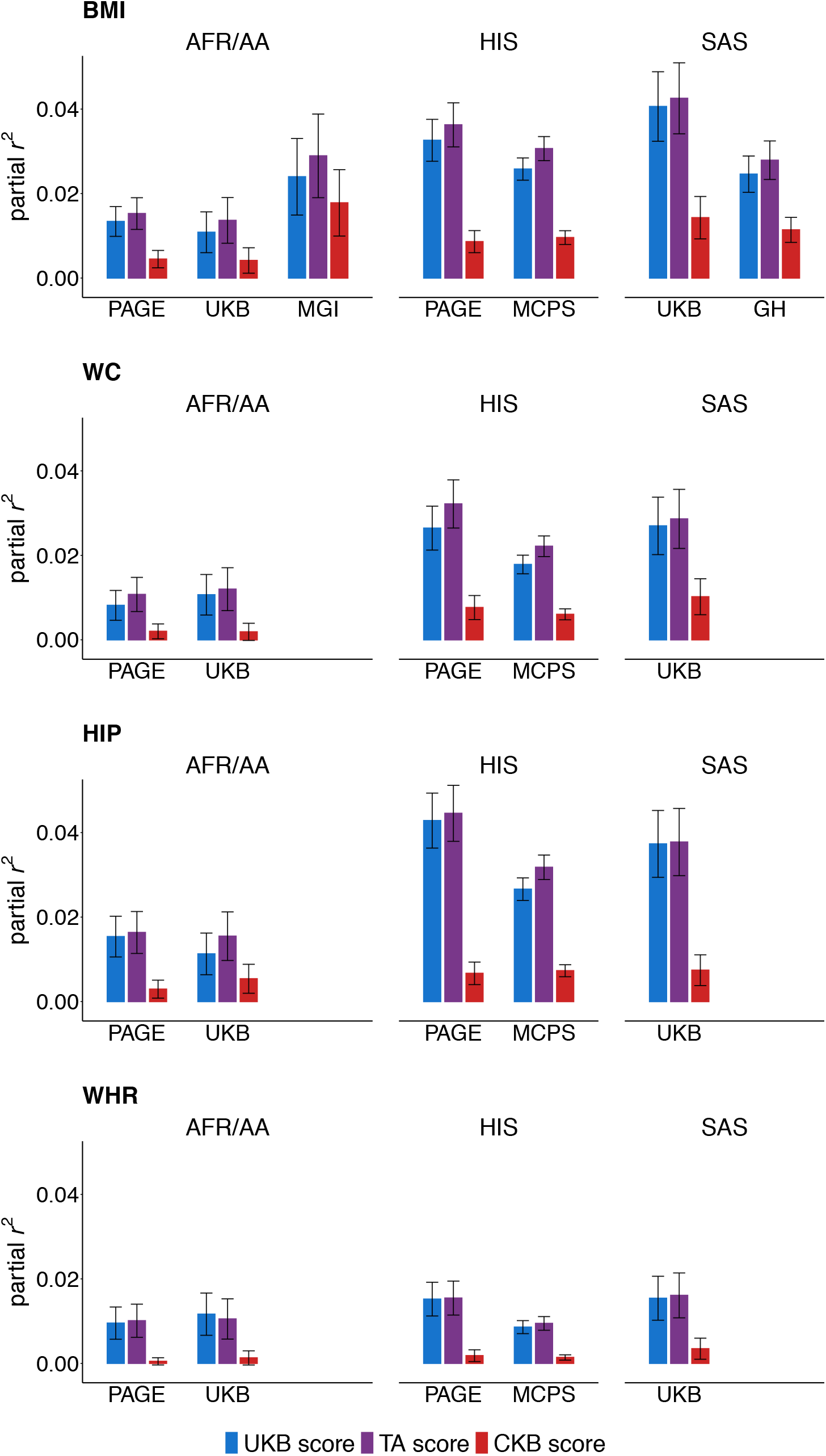
Score performance in AFR/AA, HIS and SAS populations Partial *r*^2^ values were estimated for the genetic scores in the unrelated subsets of AFR/AA, HIS and SAS populations. Values were estimated from the regression of each adiposity trait against the corresponding score with adjustment for study specific PCs and covariates, age, age^2^ and sex. The 95% CI are plotted based on the standard error of the partial *r*^2^ values estimated using Olkin and Finn’s approximation.

## Discussion

We have performed GWAS and TAMA of eight adiposity-related traits representing measures of overall adiposity or of body fat distribution, using data from two population-based cohorts of EUR and EAS ancestries. We identified 1,441 separate regions of the genome showing association with one or more of the traits. Genetic scores constructed using results from the TAMA significantly increased variance explained in multiple EAS populations over that attained using scores based on either UKB or CKB GWAS results alone.

Based on previous results from body fat imaging studies^30–32^, we classified the adiposity traits either as measures of overall adiposity or as measures of body fat distribution, which was concordant with the observational correlation of the traits in UKB and CKB (**Supplementary Figure 1**). Cross-trait genetic correlation and the quantification of locus sharing between traits showed that this distinction had a genetic underpinning. Genetic correlation between pairs of overall adiposity traits was much stronger than for other trait combinations (**Figure 2c**) and, of the 1,441 distinct regions of the genome associated with one or more of the adiposity traits in the TA results, only 50 were shared across all eight traits while 33% were associated only with one or more overall adiposity traits and another 30% only with one or more of the body fat distribution traits.

When we performed functional analysis of genes lying within loci specific to either ‘class’ of adiposity trait, we found enrichment for distinct biological processes. Regions of the genome associated only with overall adiposity showed enrichment for genes in pathways relating to neuronal processes and had upregulated expression throughout the brain (**Supplementary Table 5 and Supplementary Figure 9a**), mirroring findings from previous studies characterising BMI loci^22, 28^. In contrast, genes at loci associated with body fat distribution showed enrichment for developmental processes, and were significantly up or down regulated across a wider range of tissue types, including in: brain, subcutaneous adipose; female reproductive and circulatory tissue (**Supplementary Table 5 and Supplementary Figure 9b**). The complex tissue expression profile of genes at body fat distribution loci and their involvement in a broad range of developmental processes builds on previous reports that showed diverse pathways regulate the patterning of adipose tissue in humans^35^. However, it is important to consider that as only ∼1% of the dataset used in the functional analysis is of EAS ancestry^45^, this may limit the sensitivity of our FUMA analysis to biological pathways that are more relevant to EAS.

Although the smaller sample size of CKB gave reduced power for assessment of some aspects of the genetic architecture of adiposity traits, heritability of the body fat distribution measures was clearly lower in CKB than in UKB. Nevertheless, we found no evidence that these traits had substantial differences in the genetic architecture between EUR and EAS; trans-ancestry genetic correlation values were similar for all traits, with no clear differences between overall adiposity and fat distribution traits (**Figure 2b, Supplementary Figure 12b**).

The trait variance accounted for by genetic scores constructed based on the UKB, CKB, or TA results differed considerably between EUR and EAS populations. As is now well-established^23–26^, we found there to be substantial reductions in performance when scores derived in one ancestry were used in another (**Figure 3 and Supplementary Figure 13**). Although differences in heritability may account for some of the reduction in performance when porting scores between ancestries, in the case of the reduced performance of the UKB scores in EAS, we estimated that 42-59% can be attributed to the score’s variants having lower MAF in EAS than in EUR and to different LD patterns at the trait-associated loci. (**Figure 4a, Supplementary Table 9**). These results are consistent with a previous finding that LD and MAF differences accounted for ∼57% of the reduction in performance when transferring EUR scores to EAS for a broad range of traits and diseases^42^.

Despite only a modest increase in the number of variants included in the TA scores compared to the UKB scores, we found that in independent EAS populations the TA scores increased variance explained by up to 1.8-fold (**Figure 3**). This figure was somewhat lower than in CKB, where variance explained by the TA scores was up to 2.4-fold greater than the UKB scores (**Figure 3 and Supplementary Figure 13**). As all genotyped CKB participants were included at the discovery stage, it is not possible for us to address whether this difference in improvement is caused by overfitting in CKB or because the TAMA identified causal variants specific to the CKB population. Additionally, differences in environment between the EAS populations (e.g. altering heritability and/or affect sizes for particular variants) is also likely to be a contributing factor modifying score performance.

By comparing the performance of subsets of the UKB and TA scores which are composed of different sets of variants tagging the same regions of the genome, we found suggestive evidence that the TA scores accounted for a greater proportion of trait variation in EAS partly because variants selected in the TAMA may function as better tags of shared causal variants (**Figure 4b**). Indeed, despite seeing little benefit of the TA scores in EUR, we saw a modest but consistent improvement in variance explained by the TA scores over the UKB scores in multiple other populations of AFR/AA, SAS, and HIS ancestry (**Figure 5**). Thus, these results highlight that, although over-representation of EUR in GWAS results will likely continue to exist for many years, combining EUR GWAS data with a smaller proportion of individuals of other ancestries at the discovery stage can still lead to meaningful improvements in variance explained in non-EUR populations, even for populations of ancestries not included in the TAMA.

Since we constructed scores using only variants that reached genome-wide significance (P < 5×10^-8^), these do not perform as well as PGS based on millions of variants^10, 11^; scores constructed using PRScs^11^ based on the UKB GWAS results had variance explained in EAS cohorts approximately double that using the TA scores (**Supplementary Figure 19**). However, as our purpose was to derive genetic scores to be used for MR analyses in UKB and CKB, relaxing the criteria for variant inclusion would expose any downstream MR analysis in these datasets to overfitting and bias. Future studies may wish to utilise the summary statistics we make available from our work to construct scores for the identification of individuals at particular risk of overweight/obesity in independent datasets.

In conclusion, we assessed a wide range of adiposity measures for genetic associations using data from large EUR and EAS population-based studies and found some indication that the genetic architecture of the traits may vary between the two ancestries. By combining GWAS data from EUR and EAS and incorporating ancestry-specific LD patterns we improved variant selection, choosing conditionally independent top hits that account for a greater proportion of trait variation in EAS than variants identified from only EUR GWAS data. From this, we have developed genetic scores that account for a considerable proportion of variation in the adiposity traits in both EUR and EAS, and which have improved performance in AFR/AA, SAS and HIS populations, making them strong instruments for well powered MR analyses.

## Methods

### Sample selection

For the primary observational and genetic analyses in UKB we included 443,359 individuals of EUR descent. Criteria for individuals inclusion in the analysis were as follows: they had not withdrawn consent for their data to be used; they were of EUR ancestry; their genotypic sex matched their self-reported sex; they did not have excessive levels of heterozygosity or missingness^46^; and they had both genotype and imputation information. Individuals of EUR ancestry were identified based on both self-reporting as white British and being confirmed as genetically of EUR ancestry as described by Mahajan et al^47^. Individuals were confirmed as genetically EUR by projecting the genotyped UK Biobank samples on to the two major principal components (PCs) of four 1000 Genomes populations (CEU, YRI, CHB and JPT)^46^. Samples with PC scores falling in the neighbourhood of the CEU cluster were classified as being of EUR descent^46, 47^.

For the observational analysis in CKB we included all 513,228 participants who had not withdrawn consent. For the genetic analysis in CKB we used 96,124 genotyped individuals as previously described^48^, but excluded those individuals who were genotyped on the basis of having a COPD exacerbation due to the known impact of COPD on BMI^49^. All analysis used data from CKB release 15, accessed under Research Tracker ID: 2016-0021.

Where analyses used a subset of unrelated individuals from each dataset, in CKB 72,383 individuals were used, identified as described by Walters et al ^48^, and in UKB 366,071 individuals were identified on the basis of the relatedness coefficients (<0.05) provided with the UKB data download^46, 50^.

### Phenotype measurement and transformation

For the GWAS analyses we derived rank-inverse normal transformed traits, using values of the adiposity traits measured at baseline. To account for differences in the distribution of the traits between sexes, and across geographical regions in China, transformation was performed separately by sex in UKB and by sex and recruitment region in CKB. Prior to transformation, traits were linearly regressed against age and age^2^ (and BMI for BMI-adjusted traits) and in UKB, also against assessment centre and 40 PCs provided with the UKB data download^46, 50^. The residuals from the above regressions were then rank-inverse normal transformed. In CKB, the transformation of the traits was conducted across the full cohort used in the observational analysis.

### GWAS in UKB and CKB

GWAS for each trait was performed using BOLT-LMM v2.3.4 ^51^, calibrated using pre-computed LD scores from the EUR or EAS 1000 genomes v3 populations as supplied with the LDSC software package^40^. For each biobank the association analysis was run separately by sex using the RINT traits as input. The model was adjusted for genotyping array in both biobanks and for assessment centre in UKB. To select variants to be used in computation of the mixed model, in each biobank we filtered the imputed genotype data to retain variants with an INFO score > 0.95, MAF > 0.01 and, after conversion to best-guess genotypes using PLINK v2 (hard-call threshold 0.01), with Hardy-Weinberg P > 1×10^-6^ and missingness < 0.01. Regions of long-range LD (**Supplementary Tables 10**^52^ **and 11**^48^ for UKB and CKB respectively) were removed and the remaining list of variants was LD-pruned in the unrelated subset of each biobank using the PLINK v2 command --indep-pairwise 50 5 0.2. Analysis was run for genome build GRCh37.

### Meta-analysis

For each biobank we meta-analysed the sex-specific association statistics using the HE-RE model implemented by the software package METASOFT v3^34^. To control for inflation, we considered whether the heterogeneity inflation factors calculated by METASOFT were greater than 1. As the inflation factor was never greater than 1 no correction was applied (**Supplementary Table 6**).

We also used the HE-RE model to conduct TAMA of the 4 biobank and sex-specific sets of GWAS results for each trait. Where a variant did not have MAF > 0.01 and INFO > 0.3 in both biobanks it was still included in the TAMA but the beta and SE values were set to NA for the cohort in which the variant did not have MAF > 0.01 and INFO > 0.3. For all traits, the λ_het_ values estimated by the HE-RE model showed signs of inflation (**Supplementary Table 6**) and the relevant genomic control (GC) correction calculated by METASOFT was applied to the heterogeneity component of the HE-RE test statistic^34^ using the *-lambda_hetero* parameter.

### LDSC regression, testing for inflation, estimating heritability and cross trait-genetic correlation

Checks for potential inflation of test statistics, estimation of h^2^_SNP_ and cross-trait genetic correlation used LDSC regression applied to sex-specific and sex-combined summary statistics from each biobank. We restricted the regression to variants with chi^2^ < 30, MAF > 0.01 and INFO > 0.9, as well as removing variants lying in regions of long-range LD (**Supplementary Tables 10 and 11** for UKB and CKB respectively). We used EUR and EAS LDSCs provided with the LDSC package and the default LDSC options as documented here https://github.com/bulik/ldsc/wiki/LDSC_reg.

LDSC regression results indicated that any inflation was attributable to the polygenic nature of the traits (**Supplementary Table 6 and Supplementary Figures 2,3 and 6**), so we did not apply GC correction to the sex-specific or sex-combined summary statistics. While the overall λ_GC_ estimated for each trait from the TAMA showed signs of inflation (**Supplementary Table 6**), we again attributed this to the polygenic nature of the traits as observed for the biobank-specific test statistics, and no GC correction was applied.

To assess whether SNP-based heritability or cross-trait genetic correlation differed significantly between the biobanks, we used a two-sided test based on the normal distribution. We Bonferroni corrected the significance threshold by the number of traits, and also for the two sexes for the sex-specific comparisons. For the sex-combined heritability analysis this gave an alpha=0.006, for the sex-specific analysis alpha=0.003. For the sex-combined genetic correlation analysis this gave an alpha=0.002, for the sex-specific analysis alpha=0.0009.

### Creating LD reference sets

For use in clumping to define locus boundaries, identification of independent association signals and trans-ancestry genetic correlation analyses, we created a reference dataset from the imputed data for each biobank. Imputed dosage data from 45,000 and 15,000 unrelated study participants in UKB and CKB, respectively, were converted to best guess genotypes based on a hard call threshold of 0.49 using PLINK v2. The data were filtered to retain variants with MAF > 0.01 and INFO > 0.3 in the corresponding study population.

### Defining loci and identifying lead and secondary index signals

For each biobank, we filtered the sex-specific and sex-combined summary statistics from the GWAS of each trait, retaining only variants that were biallelic with a MAF > 0.01 and INFO > 0.3 in that dataset, and used these statistics throughout the process of defining loci and identifying variants.

To identify genomic loci associated with the adiposity traits at P < 5 × 10^−8^ in either of the biobanks, we LD clumped the sex-combined summary statistics in PLINK v1.9 using the corresponding reference dataset as an LD reference panel. We set the clumping algorithm to identify top-associated variants with a P < 5 × 10^−8^ and all variants within a ±5 mega base (MB) window of the top associated variant, that were in LD with the top variant (r^2^ > 0.05) and had an association P-value < 0.05. We then added ±1000 bp to the LD based clumps and, when clumps overlapped in terms of genomic span, they were merged. The genomic span of the LD based clumps defined the bp position of loci associated with the adiposity traits in either biobank. We took the most significantly associated variant within a locus to be the locus’s lead variant.

To identify secondary signals at the loci defined in either the UKB or CKB analysis, we took an iterative approach involving rounds of conditional analysis at each locus separately using the Genome-wide Complex Trait Analysis (GCTA)^53^ software package and then meta-analysis using METASOFT. See **Supplementary Figure 4** for a schematic of the conditional analysis process.

In the first stage of the pipeline, we conditioned the relationship between all variants at a locus and the corresponding adiposity trait on the effect of the locus’ lead variant by implementing the GCTA *--cojo-cond* command using the sex-specific summary statistics as input. We then meta-analysed the output from the parallel sex-specific conditional analyses using the HE-RE model, to give sex-combined association statistics for the locus conditioned on the lead variant.

If no variants at the locus were significantly associated (HE-RE P-value < 5×10^−8^) with the corresponding trait in the meta-analysed conditional results, no further conditional analysis was performed on the region. When variants within the locus continued to be associated, we fit the most significant variant in a joint conditional model with the lead variant using the GCTA command *--cojo-joint* and meta-analysed the sex-specific output using the HE-RE model.

When both variants fitted in the joint model remained significant (P < 5×10^-8^) in the meta-analysed results the pipeline was started again, this time conditioning the locus on the effect of the lead variant and the newly detected ‘secondary signal’. If one or more variants were not significant when fit together in the joint model, we excluded the non-significant variant(s) and reconditioned the locus on the effect of the remaining significant variant(s) using the same method (GCTA command *--cojo-cond*) as already detailed and continued through the pipeline from that stage onwards.

This pipeline was repeated for each locus associated with each trait in each biobank until no additional variants at any of the loci were independently associated (P < 5×10^−8^) with the corresponding trait in the meta-analysed conditional results.

### Defining loci and identifying index and secondary signals from TA results

The TA summary statistics were clumped using the same method and criteria as detailed for the biobank specific analyses, but the process was done twice, separately using the UKB and CKB reference panels. Where clumps specified using either the UKB or CKB reference set overlapped they were merged to define the bp boundaries of the TA loci (**Supplementary Figure 5**).

When defining the lead variant at a locus, we prioritised variants that were genome-wide significant and had information included in the TAMA from both UKB and CKB, even where variants that only had association statistics included from one of the biobanks were more significant. To identify secondary signals at the loci we used the same pipeline described above for the biobank-specific analyses, but the conditional analysis at each locus was performed using the separate biobank and sex-specific summary stats and the separate biobank-specific LD reference panels (**Supplementary Figure 4**). When meta-analysing the conditioned results using the HE-RE model we additionally applied the correction for the heterogeneity inflation factor estimated previously for each trait when the genome-wide GWAS summary statistics were TA meta-analysed.

### Locus novelty

For a given adiposity trait, we defined loci as novel based on whether there were any genome-wide significant associations previously reported in the same region of the genome in the GWAS Catalog or in results published by the GIANT consortium^20–22, 35–38^. To assess this we downloaded association results from https://www.ebi.ac.uk/gwas_catalog (accessed on 27/03/2022) for the adiposity traits using the search terms: “body mass index”, “body fat percentage”, “waist circumference”, “BMI-adjusted waist circumference”, “hip circumference”, “BMI-adjusted hip circumference”, “waist-hip ratio” and “BMI-adjusted waist-hip ratio” and also extracted the lead variants reported in GIANT publications for the adiposity traits. We considered loci identified in the UKB/CKB/TA analyses as novel if their bp boundaries did not overlap with 500 kb up and downstream of variants that were associated with the corresponding trait at P < 5×10^-8^ in the GWAS catalog data or GIANT results.

### Gene-set enrichment and tissue expression profiling

Gene-set enrichment and tissue expression profiles used the FUMA GENE2FUNC module^39^ applied to genes located within 10kb of index variants associated with overall adiposity or body fat distribution traits. To compare the gene-sets that genes lying in proximity to variants at overall adiposity or body fat distribution loci were enriched for, we searched the GO biological processes terms for strings relating to neuronal processes (“brain, neuro, axon, nervous, synap, cognition”) or developmental processes (“develop, morpho, embryo, differen”).

We also performed gene-set enrichment and tissue expression profiling separately for each adiposity trait, using lists of genes that were located within 10kb of the index variants associated with each trait.

### Trans-ancestry genetic correlation

We estimated the trans-ancestry genetic correlation for each adiposity trait using the software program Popcorn^41^. Using the command ‘popcorn compute -v 1’, we first computed cross-population scores from our biobank specific LD reference sets, restricting the analysis to variants with MAF > 0.01 in both datasets. These scores and the GWAS summary statistics, filtered using the same criteria as for LDSC analyses, were then used for estimation of the trans-ancestry genetic correlation values. To asses if *r*_g_ was significantly less than 1, we used a Bonferroni correction to account for the number of traits assessed, (alpha=0.006), and also for the two sexes in the sex-specific comparisons (alpha=0.003).

### Constructing and testing scores

Genetic scores for a trait were composed of conditionally independent variants identified in either the corresponding UKB, CKB or TA results. To weight the scores, we either used effect sizes estimated from our conditional pipeline (see above, and **Supplementary data tables 1-3**) or, where the weights were to be applied to the same dataset in which they were estimated, we derived weights using block jack-knifing (see **Supplementary Table 12** for the weighting of the scores used in the different datasets). For the jack-knifing, unrelated subsets of UKB or CKB were split into 100 groups, which were iteratively removed from a multivariable linear regression model of each RINT adiposity trait against all the corresponding variants for that score type (i.e. UKB/CKB/TA) with adjustment for 40 (UKB) or 12 (CKB) PCs. For the group of individuals excluded from a given regression we derived the given score as the sum of the trait increasing alleles weighted by the effect sizes estimated in the 99% of unrelated individuals who were included in the regression. In the subset of each biobank excluded on the basis of relatedness, we derived the scores as the sum of the trait increasing alleles multiplied by the effect sizes estimated from multivariate regressions fitted in the total unrelated dataset of each biobank.

We assessed the performance of the scores in each biobank by calculating partial *r*^2^ values. To do this, we fit one linear model regressing each adiposity trait against age, age^2^, sex and 40 and 12 PCs in UKB and CKB, respectively, and a second model that also included either the corresponding UKB, CKB or TA score. We then estimated the partial *r*^2^ value as the ratio of the difference between the error sum of squares of the reduced model and the full model to the error sum of squares of the reduced model. We estimated the standard error of the partial *r*^2^ values using Olkin and Finn’s approximation^54^.

### Testing scores in external data

We tested the UKB, CKB and TA scores in the external studies: EPIC-Norfolk (EPIC)^55^; The Fenland Study (FS)^56^; Michigan Genomics Initiative (MGI)^57^; The Trφndelag Health Study (HUNT)^58, 59^; Oxford Biobank (OXBB)^60^; The Avon Longitudinal Study of Parents and Children (ALSPAC)^61^; Taiwan Biobank (TWB)^62^; China Health and Nutrition Survey (CNHS)^63^; BioBank Japan (BBJ)^64^; The Population Architecture using Genomics and Epidemiology study (PAGE) ^43^; Mexico City Prospective Study (MCPS)^44^ and East London Genes and Health (GH)^65^.

We also tested the scores for individuals of AFR or SAS descent in UKB, who were not included in the primary UKB analysis. In UKB, we defined individuals as being of SAS or AFR ancestry based on individuals self-reporting as having SAS or AFR ancestry and following PCA falling within 5 SDs of the 1000 genome SAS and AFR populations.

Partial r^2^ values were estimated in the external datasets using the same method as applied in UKB and CKB, for the weighting of the scores see Supplementary Table 12. Adjustments made in the linear regression were study specific, for the adjustments made by each study see **Supplementary Table 13**. Some studies (OXBB, MGI, HUNT and ALSPAC) had 15% or more of variants from the UKB/CKB/TA scores missing from their datasets, so, to maximise comparability between studies, we did not include results from these studies in our main analyses. See **Supplementary Figure 20** for results from these datasets.

The assessment of the scores in MCPS stratified by proportion of EUR ancestry was informed by ADMIXTURE analysis in the cohort, detailed by Ziyatdinov et al^66^.

### Investigating factors affecting the transferability of genetic risk scores

To approximate the contribution of MAF and LD differences between EUR and EAS to the reduced relative performance of the UKB genetic scores in CKB, we used a formula devised by Wang et al^42^. The formula requires LD independent variants so we LD-clumped the conditionally independent variants identified in the UKB analysis using an *r*^2^ threshold of 0.01 in a window size of 1Mb. As input to the formula we used EUR and EAS whole genome sequence data from the 1000 genomes v3 to estimate LD between each UKB LD-independent variant and all candidate causal variants (variants lying in a 100kb window around each UKB variant that were also in LD *r*^2^ > 0.45 with it). We additionally used as input the MAF of the UKB LD-independent variants in UKB and CKB and the UKB effect sizes. We re-constructed the UKB scores based only on the LD-independent variant set and tested them in CKB and EPIC.

## Data Availability

CKB phenotype data are available under the CKB Open Access Data Policy to bona fide researchers. Sharing of genotyping data is currently constrained by the Administrative Regulations on Human Genetic Resources of the People’s Republic of China. Access to these and certain other data is available through collaboration with CKB researchers. Full details of the CKB Data Sharing Policy are available at www.ckbiobank.org. The UKB genotype and phenotype data were accessed under project numbers 50474 and 9161 (http://www.ukbiobank.ac.uk/). UKB data can be accessed upon request once a research project has been submitted and approved by the UKB committee.

## Acknowledgements

The chief acknowledgment is to the CKB and UKB participants. We also thank the CKB project staff, and the China National Centre for Disease Control and Prevention (CDC) and its regional offices for assisting with the fieldwork

The CKB baseline survey and the first re-survey were supported by the Kadoorie Charitable Foundation in Hong Kong. The long-term follow-up has been supported by Wellcome grants to Oxford University (212946/Z/18/Z, 202922/Z/16/Z, 104085/Z/14/Z, 088158/Z/09/Z) and grants from the National Key Research and Development Program of China (2016YFC0900500, 2016YFC0900501, 2016YFC0900504, 2016YFC1303904) and from the National Natural Science Foundation of China (91843302). DNA extraction and genotyping was supported by grants from GlaxoSmithKline and the UK Medical Research Council (MC-PC-13049, MC-PC-14135). The UK Medical Research Council (MC_UU_00017/1,MC_UU_12026/2 MC_U137686851), Cancer Research UK (C16077/A29186; C500/A16896) and the British Heart Foundation (CH/1996001/9454) provide core funding to the Clinical Trial Service Unit and Epidemiological Studies Unit at Oxford University for the project.

ZFH was partly funded by a grant from The Wellcome Trust (H5R00150). The computational aspects of this research were supported by the Wellcome Trust Core Award Grant Number 203141/Z/16/Z and the NIHR Oxford BRC. The views expressed are those of the author(s) and not necessarily those of the NHS, the NIHR or the Department of Health.

YFL. is supported by the National Health Research Institutes (NP-110-PP-09; NP-111-PP-09) and the Ministry of Science and Technology (110-2314-B-400-028-MY3) of Taiwan. We thank the National Core Facility for Biopharmaceuticals (NCFB, MOST 106-2319-B-492-002) and National Center for High-performance Computing (NCHC) of National Applied Research Laboratories (NARLabs) of Taiwan for providing computational and storage resources.

The Mexico City Prospective Study has received funding from the Mexican Health Ministry, the National Council of Science and Technology for Mexico, the Wellcome Trust, Cancer Research UK, British Heart Foundation and the UK Medical Research Council.

GH is funded by The Wellcome Trust and Royal Society (208806/Z/17/Z). The UK Medical Research Council and Wellcome (217065/Z/19/Z and WT088806) and the University of Bristol provide core support for ALSPAC. This publication is the work of the authors and RGW and GH will serve as guarantor for the contents of this paper.

The BioBank Japan Project was supported by AMED under Grant Number JP21km0605001.

LB and BB receive support from the K.G. Jebsen Center for Genetic Epidemiology funded by Stiftelsen Kristian Gerhard Jebsen; Faculty of Medicine and Health Sciences, NTNU; The Liaison Committee for education, research and innovation in Central Norway; and the Joint Research Committee between St Olavs Hospital and the Faculty of Medicine and Health Sciences, NTNU.

The Population Architecture Using Genomics and Epidemiology (PAGE) program is funded by the National Human Genome Research Institute (NHGRI) with co-funding from the National Institute on Minority Health and Health Disparities (NIMHD). The contents of this paper are solely the responsibility of the authors and do not necessarily represent the official views of the NIH. The PAGE consortium thanks the staff and participants of all PAGE studies for their important contributions. We thank Rasheeda Williams and Margaret Ginoza for providing assistance with program coordination. The complete list of PAGE members can be found at http://www.pagestudy.org.

Assistance with data management, data integration, data dissemination, genotype imputation, ancestry deconvolution, population genetics, analysis pipelines, and general study coordination was provided by the PAGE Coordinating Center (NIH U01HG007419). Genotyping services were provided by the Center for Inherited Disease Research (CIDR). CIDR is fully funded through a federal contract from the National Institutes of Health to The Johns Hopkins University, contract number HHSN268201200008I. Genotype data quality control and quality assurance services were provided by the Genetic Analysis Center in the Biostatistics Department of the University of Washington, through support provided by the CIDR contract.

The data and materials included in this report result from collaboration between the following studies and organizations:

BioMe Biobank: Samples and data of The Charles Bronfman Institute for Personalized Medicine (IPM) BioMe Biobank used in this study were provided by The Charles Bronfman Institute for Personalized Medicine at the Icahn School of Medicine at Mount Sinai (New York). Phenotype data collection was supported by The Andrea and Charles Bronfman Philanthropies. Funding support for the Population Architecture Using Genomics and Epidemiology (PAGE) IPM BioMe Biobank study was provided through the National Human Genome Research Institute (NIH U01HG007417).

HCHS/SOL: Primary funding support to Dr. North and colleagues is provided by U01HG007416. Additional support was provided via R01DK101855 and 15GRNT25880008. The Hispanic Community Health Study/Study of Latinos was carried out as a collaborative study supported by contracts from the National Heart, Lung, and Blood Institute (NHLBI) to the University of North Carolina (N01-HC65233), University of Miami (N01-HC65234), Albert Einstein College of Medicine (N01-HC65235), Northwestern University (N01-HC65236), and San Diego State University (N01-HC65237). The following Institutes/Centers/Offices contribute to the HCHS/SOL through a transfer of funds to the NHLBI: National Institute on Minority Health and Health Disparities, National Institute on Deafness and Other Communication Disorders, National Institute of Dental and Craniofacial Research, National Institute of Diabetes and Digestive and Kidney Diseases, National Institute of Neurological Disorders and Stroke, NIH Institution-Office of Dietary Supplements.

MEC: The Multiethnic Cohort study (MEC) characterization of epidemiological architecture is funded through the NHGRI Population Architecture Using Genomics and Epidemiology (PAGE) program (NIH U01 HG007397). The MEC study is funded through the National Cancer Institute U01 CA164973.

PAGE Global Reference Panel: The Stanford Global Reference Panel was created by Stanford-contributed samples and comprises multiple datasets from multiple researchers across the world designed to provide a resource for any researchers interested in diverse population data on the Multi-Ethnic Global Array (MEGA), funded by the NHGRI PAGE program (NIH U01HG007419). The authors thank the researchers and research participants who made this dataset available to the community. The specific datasets are:

Mexico: Samples of indigenous origin in Oaxaca were kindly provided by Drs. Karla Sandoval Mendoza, Samuel Canizales Quinteros, and Victor Acuña Alonzo. Peru: Individuals from a primarily Quechuan and Aymaran-speaking community in Puno, Peru were kindly provided by Drs. Julie Baker and Carlos Bustamante, with funding support from the Burroughs Welcome Fund. Rapa Nui (Easter Island): Samples were kindly provided by Drs. Karla Sandoval Mendoza and Andres Moreno Estrada with funding from the Charles Rosenkranz Prize for Health Care Research in Developing Countries.

South Africa: Samples of KhoeSan individuals from the ‡Khomani and Nama communities were kindly provided by Drs. Brenna Henn and Christopher Gignoux with funding from the Morrison Institute for Population and Resource Studies. Honduras and Colombia: Samples from communities in Honduras and Colombia were kindly provided by Dr. Kathleen Barnes (University of Colorado, Denver), Edwin Herraro-Paz (Universidad Católica de Honduras, San Pedro Sula, Honduras), Alvaro Mayorga (Universidad Católica de Honduras, San Pedro Sula, Honduras), Luis Caraballo (University of Cartagena), Javier Marrugo (university of Cartagena) Additional global samples: The following datasets are open access and available through the lab website of Carlos Bustamante (https://bustamantelab.stanford.edu/). The Human Genome Diversity Panel (HGDP-CEPH) is a group of cell lines maintained by the Centre d’Étude du Polymorphisme Humain, Fondation Jean Dausset (Paris, France) comprising 52 diverse populations across the world (Africa, Near East, Europe, South Asia, Central Asia, East Asia, Oceania and the Americas). Additional information on these datasets can be found on the CEPH website (http://www.cephb.fr/en/hgdp_panel.php), or originally at http://www.ncbi.nlm.nih.gov/pubmed/11954565 and http://www.ncbi.nlm.nih.gov/pubmed/12493913, with numerous subsequent publications. Samples were filtered to include the H952 unrelated individuals as published here: http://www.ncbi.nlm.nih.gov/pubmed/17044859. Also available on the Bustamante Lab website is genotype data for the Maasai from Kinyawa, Kenya (MKK) samples maintained by the Coriell Institute for Medical Research (https://catalog.coriell.org/1/NHGRI/Collections/HapMap-Collections/Maasai-in-Kinyawa-Kenya-MKK) and genotyped as part of the International HapMap Project Phase 3(http://hapmap.ncbi.nlm.nih.gov/, http://www.sanger.ac.uk/resources/downloads/human/hapmap3.html). We have genotyped a subset of unrelated individuals using the filters recommended in http://www.ncbi.nlm.nih.gov/pubmed/20869033.

WHI: Funding support for the “Exonic variants and their relation to complex traits in minorities of the WHI” study is provided through the NHGRI PAGE program (NIH U01HG007376). The WHI program is funded by the National Heart, Lung, and Blood Institute, National Institutes of Health, U.S. Department of Health and Human Services through contracts HHSN268201100046C, HHSN268201100001C, HHSN268201100002C, HHSN268201100003C, HHSN268201100004C, and HHSN271201100004C. The authors thank the WHI investigators and staff for their dedication, and the study participants for making the program possible. A listing of WHI investigators can be found at: https://www.whi.org/researchers/Documents%20%20Write%20a%20Paper/WHI%20Investigator%20Short%20List.pdf

## Authors’ contributions

ZFH analysed the data and drafted the manuscript. ZFH, MVH, MMcC and RGW contributed to the conception of this paper. ZFH, MVH, IYM, ZC, and RGW contributed to the interpretation of the results and the revision of manuscript. CK, AP, KL, WG, and AM contributed to analytical discussions. LL, ZC, IYM, CK, YC, YG, HD, KL, SS, CY, JLv, and RGW contributed to data acquisition. MJN, FK, YFL, TTC, GH, LM, BB, SG, CJW, JMT, PKM, RTC, JAD, MK, YK, MG, KEN, RJFL, CAH, UP, SB, CRG, GW, KK, SF, XM, JL and MK contributed to the replication analysis. All authors critically reviewed the manuscript and approved the final submission.

## Additional information

Supplementary Information is available for this paper.

Correspondence and requests for materials should be addressed to RGW

## Abbreviations

AFR/AA: African/African American

BBJ: Biobank Japan

BMI: body mass index

bp: base pair

CKB: China Kadoorie Biobank

CHNS: China Health and Nutrition Survey

DEG: differentially expressed genes

EAS: East Asian

EUR: European

Fat%: body fat percentage

FDR: false discovery rate

GCTA: Genome-wide Complex Trait Analysis

GWAS: genome-wide association study

HE-RE: Han-Eskin random effects

HIP: hip circumference

HIPadjBMI: HIP adjusted for BMI

HIS: Hispanic

kb: kilobase pair

LD: linkage disequilibrium

LDSC: LD score

MAF: minor allele frequency

MR: Mendelian randomisation

PGS: polygenic score

SAS: South Asian

TA: trans-ancestry

TAMA: trans-ancestry meta-analysis

TWB: Taiwan Biobank

WC: waist circumference

WCadjBMI: WC adjusted for body mass index

UKB: UK Biobank

WHR: waist-hip ratio

WHRadjBMI: WHR adjusted for BMI

