## Supplementary Figures for "Trans-ancestry meta-analysis improves performance of genetic scores for multiple adiposity-related traits in East Asian populations"

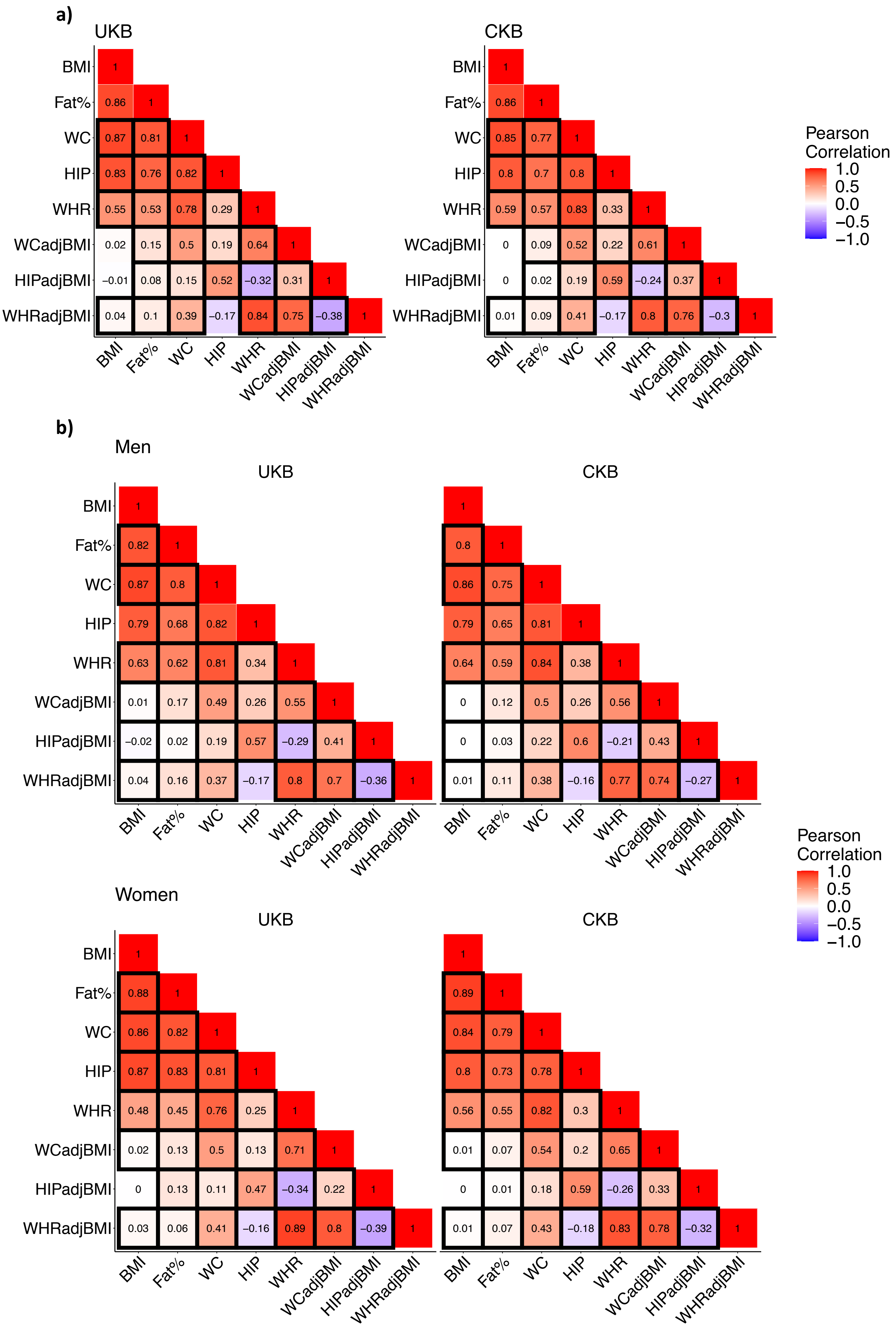


**S Figure 1) Pearson's correlation of the adiposity traits in UKB and CKB**

a) Pearson's correlation between the traits estimated for sexes combined. We used a two-sided test based on the normal distribution to compare *r* between UKB and CKB, black boxes indicate *r* is significantly different between the datasets (P-values $<10^{-6}$). The alpha threshold was Bonferroni corrected by the 28 cross-trait comparisons made.

b) Pearson's correlation estimated stratified by sex, black boxes indicate *r* is significantly different between the biobanks for each sex (P-values $<10^{-3}$). The alpha threshold was Bonferroni corrected by the 28 cross-trait comparisons made for the 2 sexes.

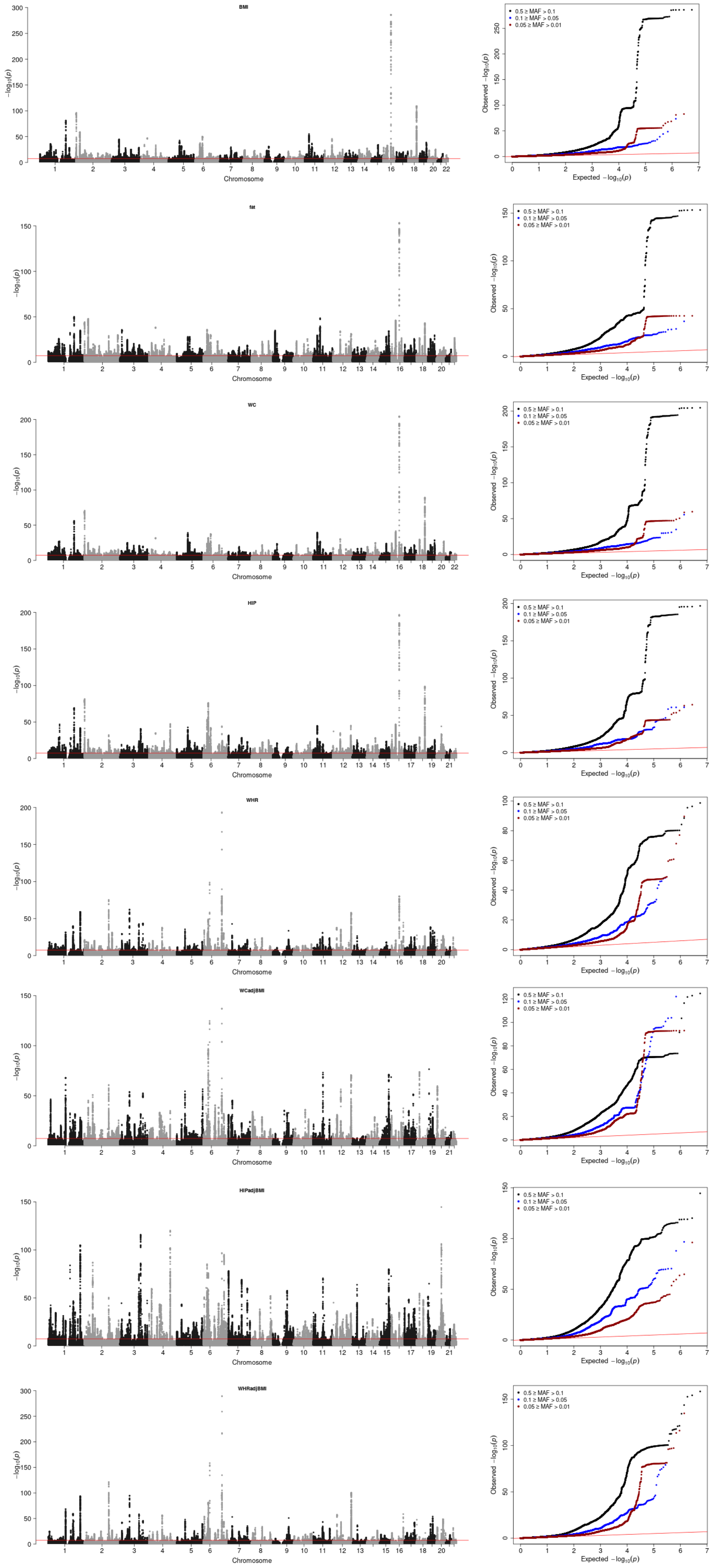


**S Figure 2) Manhattan plots and MAF-stratified QQ-plots for UKB GWAS results**

In the Manhattan plots, only association P-values < 0.05 are plotted. The significance threshold (P = 5 × 10−8) for association with the adiposity traits is plotted vertically in red. QQ plots are stratified by variants MAF in UKB

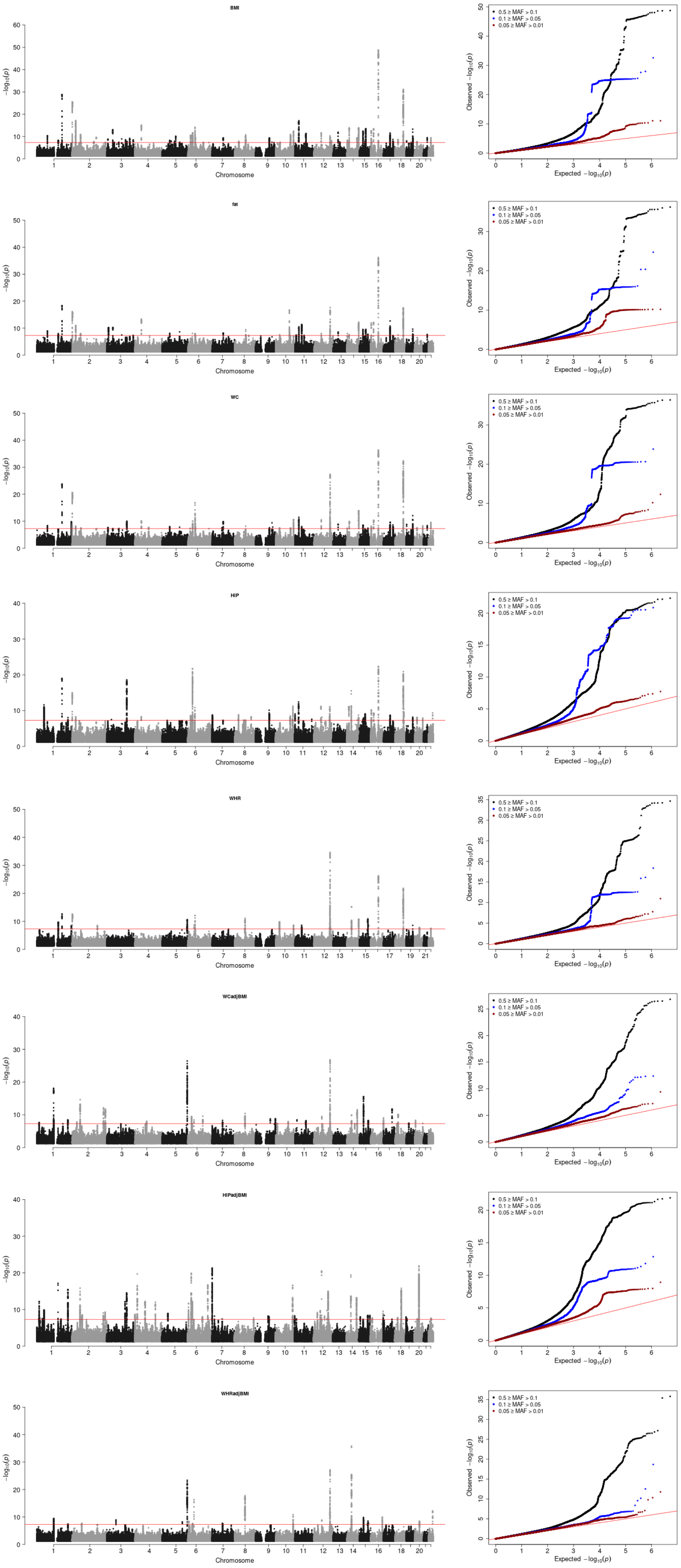


**S Figure 3) Manhattan plots and MAF-stratified QQ-plots for CKB GWAS results**

In the Manhattan plots, only association P-values < 0.05 are plotted. The significance threshold (P = 5 × 10−8) for association with the adiposity traits is plotted vertically in red. QQ plots are stratified by variants MAF in CKB.

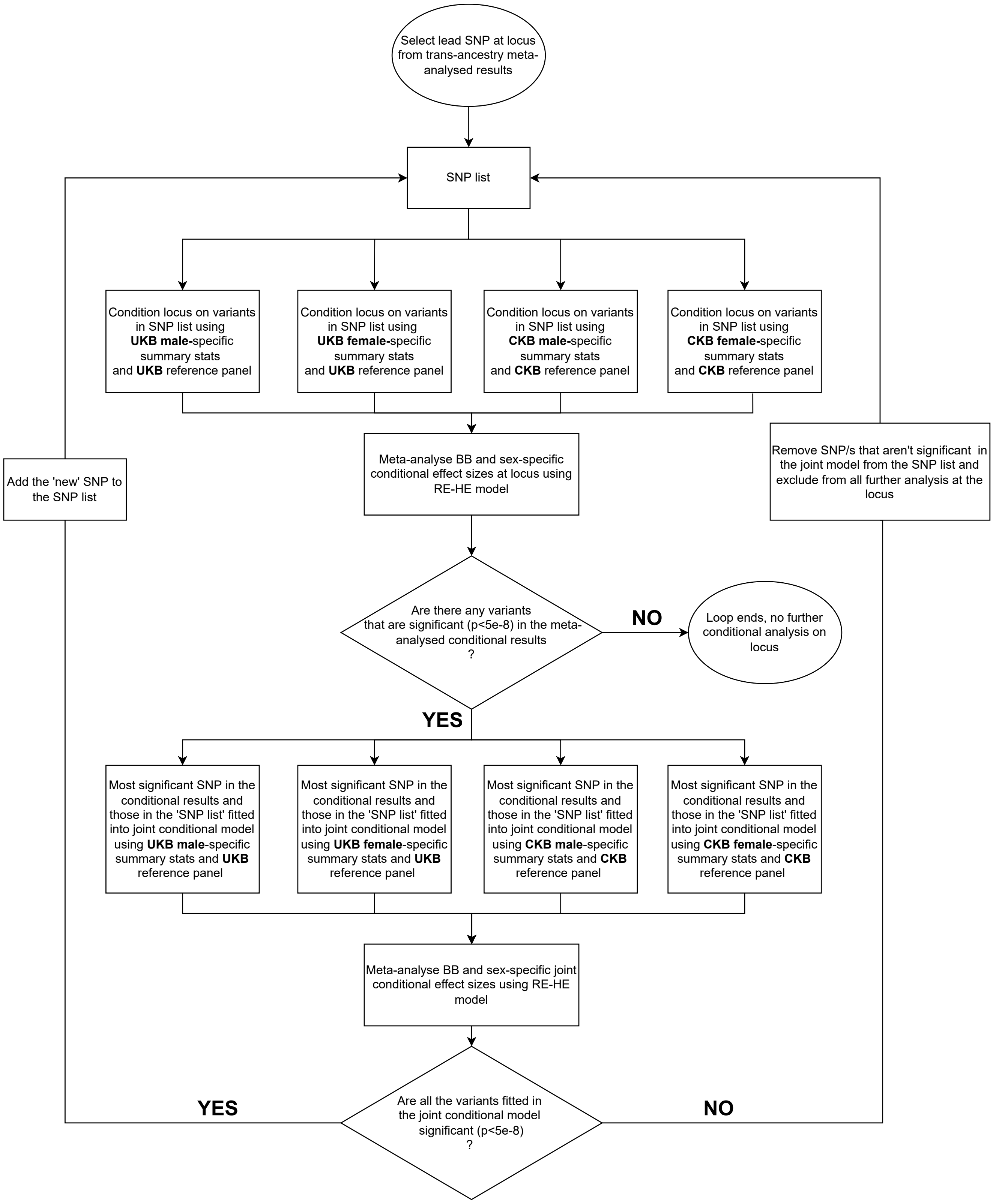


**S Figure 4) Flow diagram of the pipeline used to identify index variants from the TAMA results**

To identify TA index variants we developed an iterative approach to conditional analysis using LD reference sets from both UKB and CKB to estimate conditional effect sizes at each TA loci, followed by TAMA of the biobank (BB) and sex-specific conditional results.

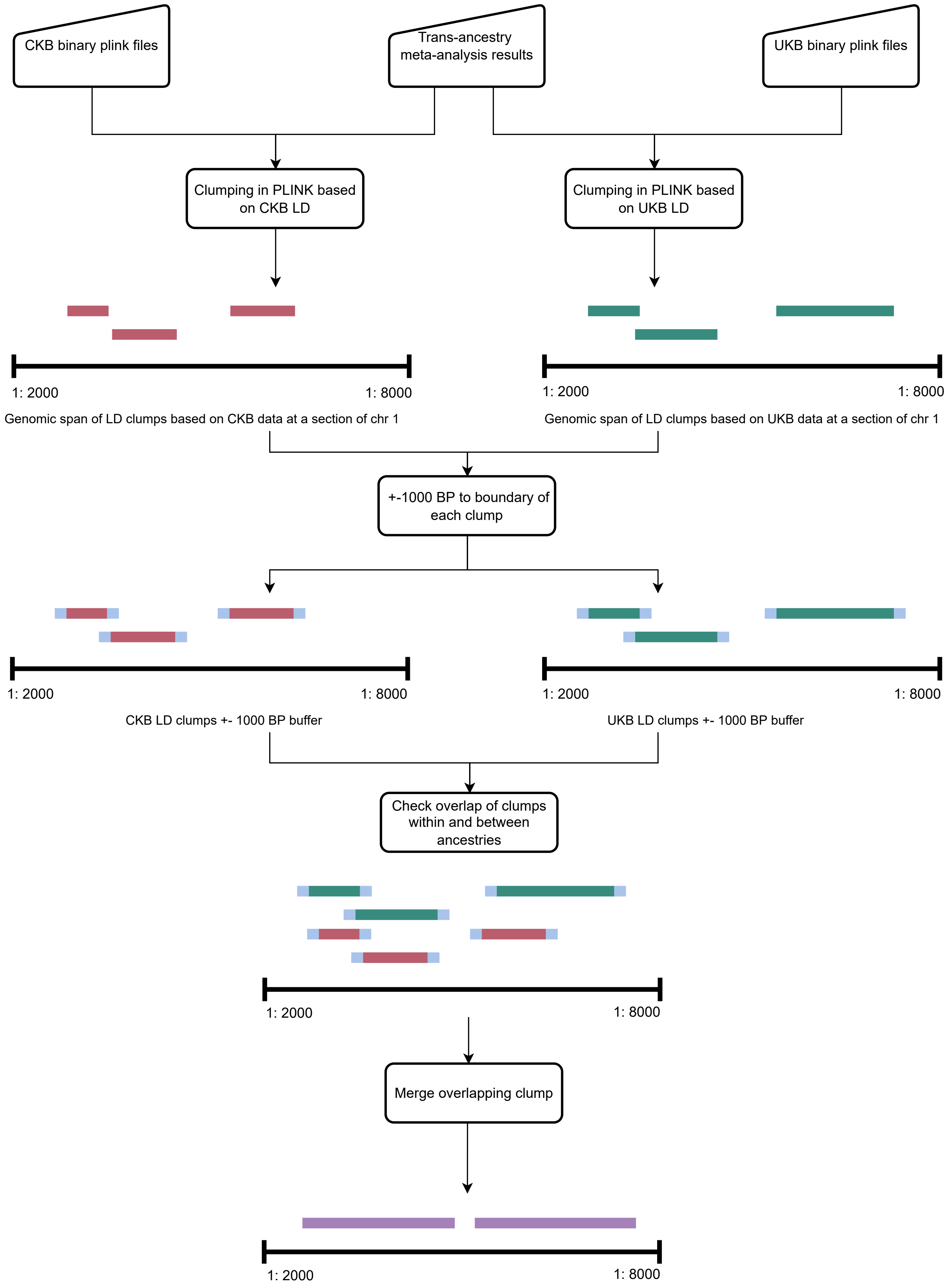


**S Figure 5) Clumping process used to define loci in the TAMA**
To define TA loci we used LD reference sets from both UKB and CKB to LD clump the TAMA results. We added 1000 BP up and downstream to the ancestry 'specific' clumps and where they overlapped we merged them into single clumps.

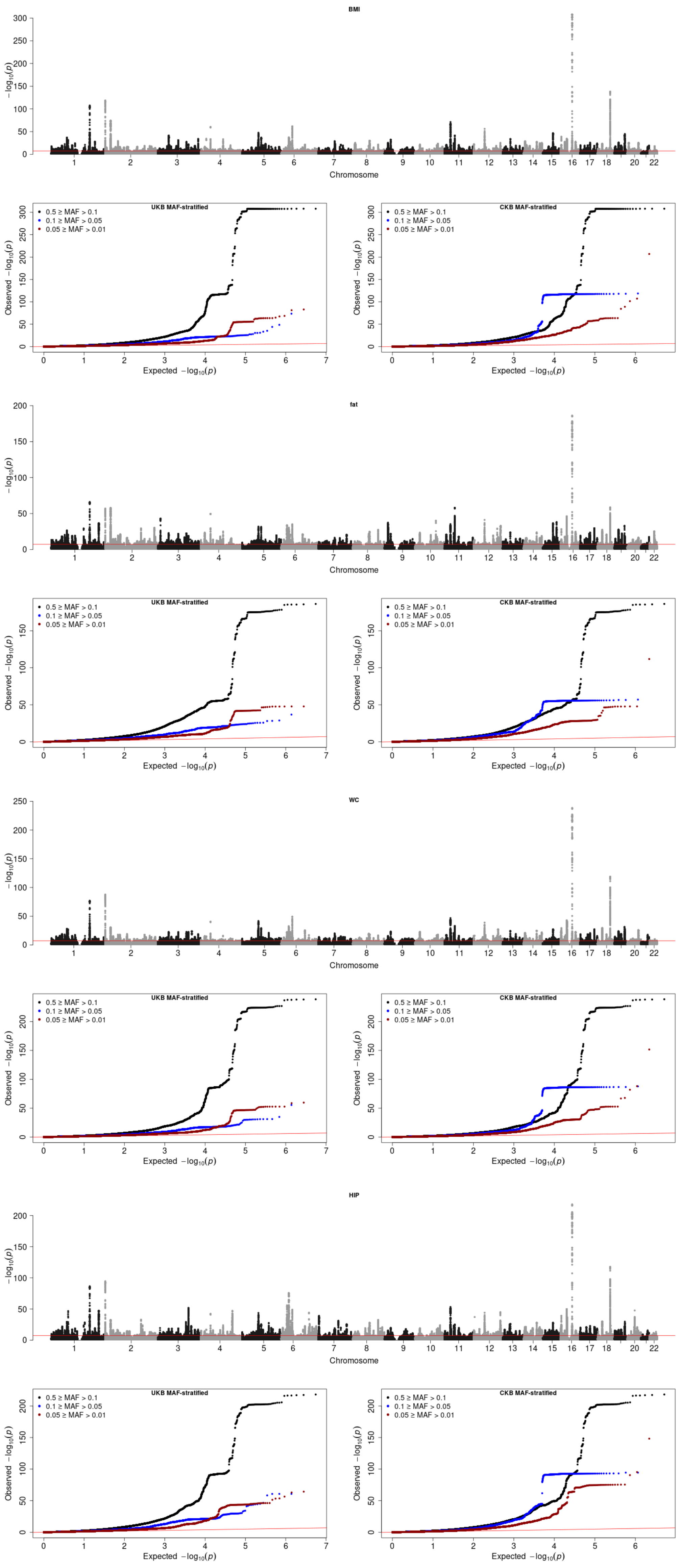


**S Figure 6a) Manhattan plots and MAF-stratified QQ-plots for TAMA results**

In the Manhattan plots, only association P-values < 0.05 are plotted. The significance threshold (P = 5 × 10−8) for association with the adiposity traits is plotted vertically in red. QQ plots are stratified by variants MAF in UKB and also in CKB.

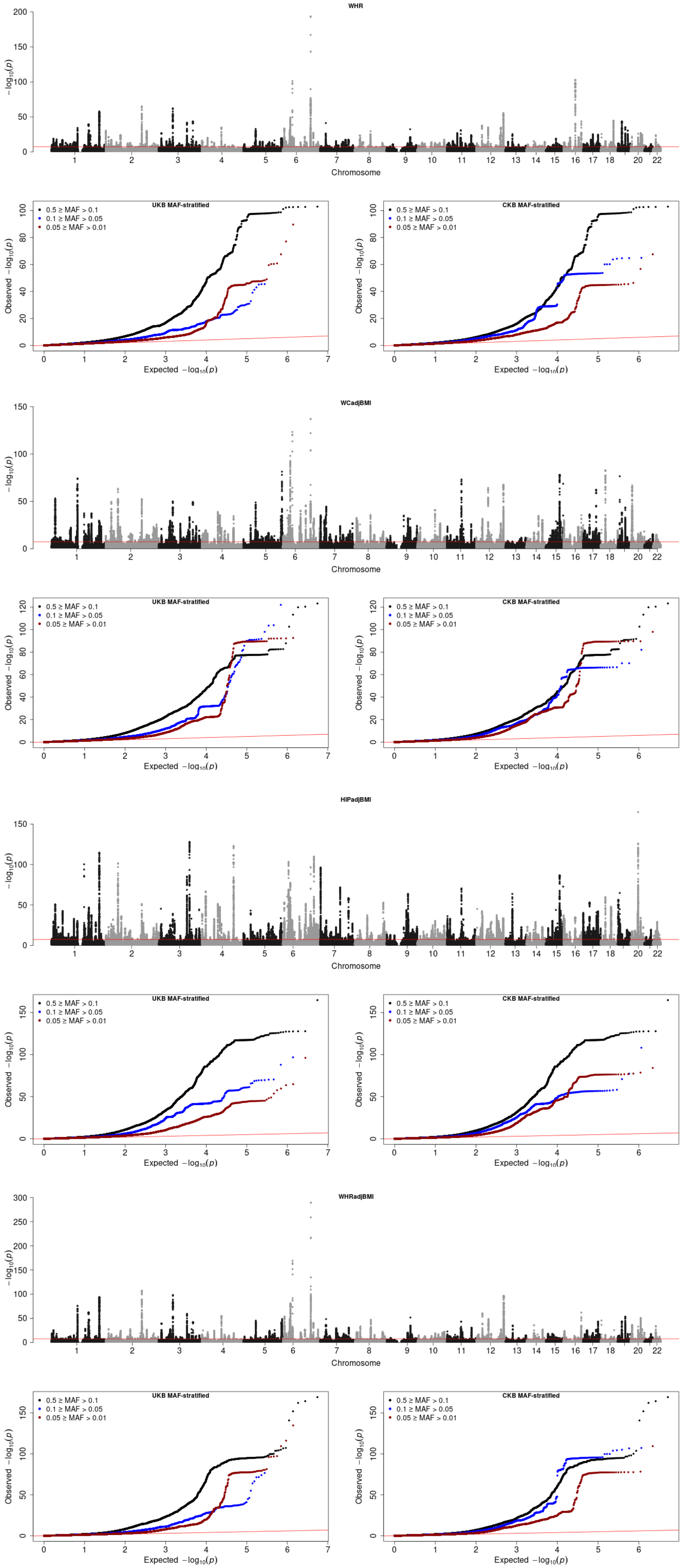


**S Figure 6b) Manhattan plots and MAF-stratified QQ-plots for TAMA results**

In the Manhattan plots, only association P-values < 0.05 are plotted. The significance threshold (P = 5 × 10−8) for association with the adiposity traits is plotted vertically in red. QQ plots are stratified by variants MAF in UKB and also in CKB.

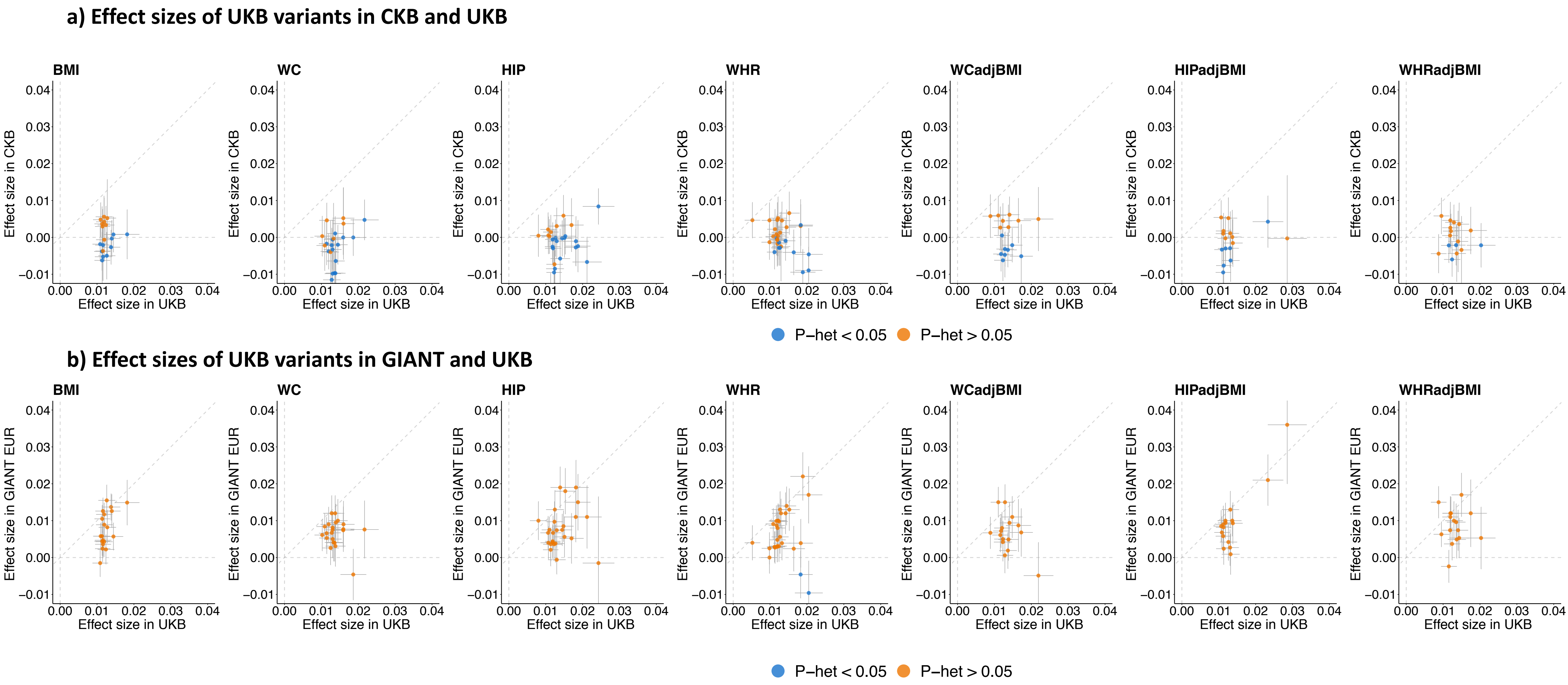


**S Figure 7) Effect sizes of variants at loci identified in the UKB analysis and not in the TAMA**

Effect sizes are reported on a SD scale, error bars represent the standard error of the estimate. Associations that show some evidence ($P_{het}$ adjusted for false discovery rate (FDR) < 0.05) for heterogeneity in effect between CKB and UKB or GIANT and UKB are highlighted in blue.

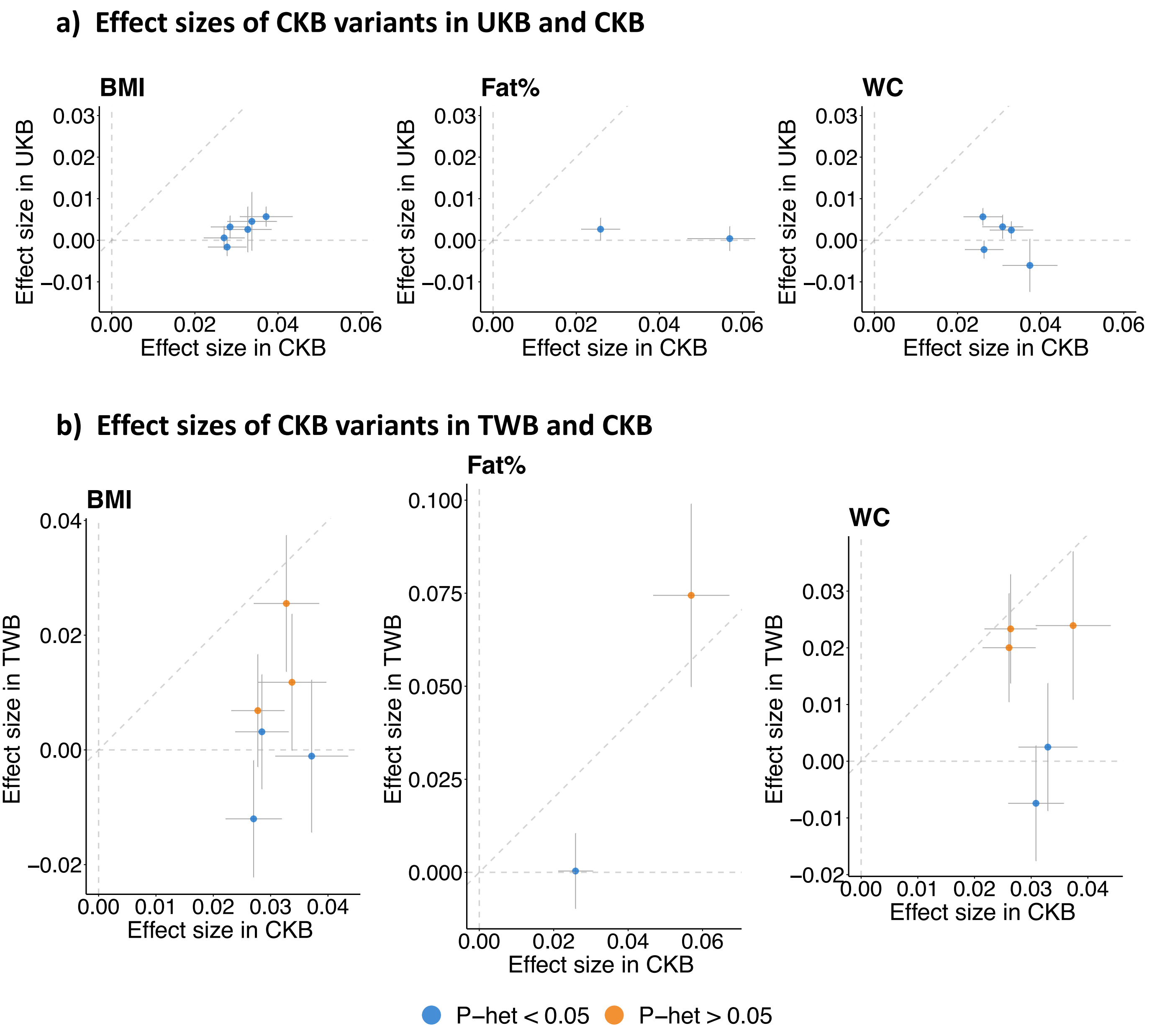


**S Figure 8) Effect sizes of variants at loci identified in the CKB analysis and not in the TAMA**
Effect sizes are reported on a SD scale, error bars represent the standard error of the estimate. Associations that show some evidence ($P_{het}$ adjusted for FDR < 0.05) for heterogeneity in effect between UKB and CKB or TWB and CKB are highlighted in blue.

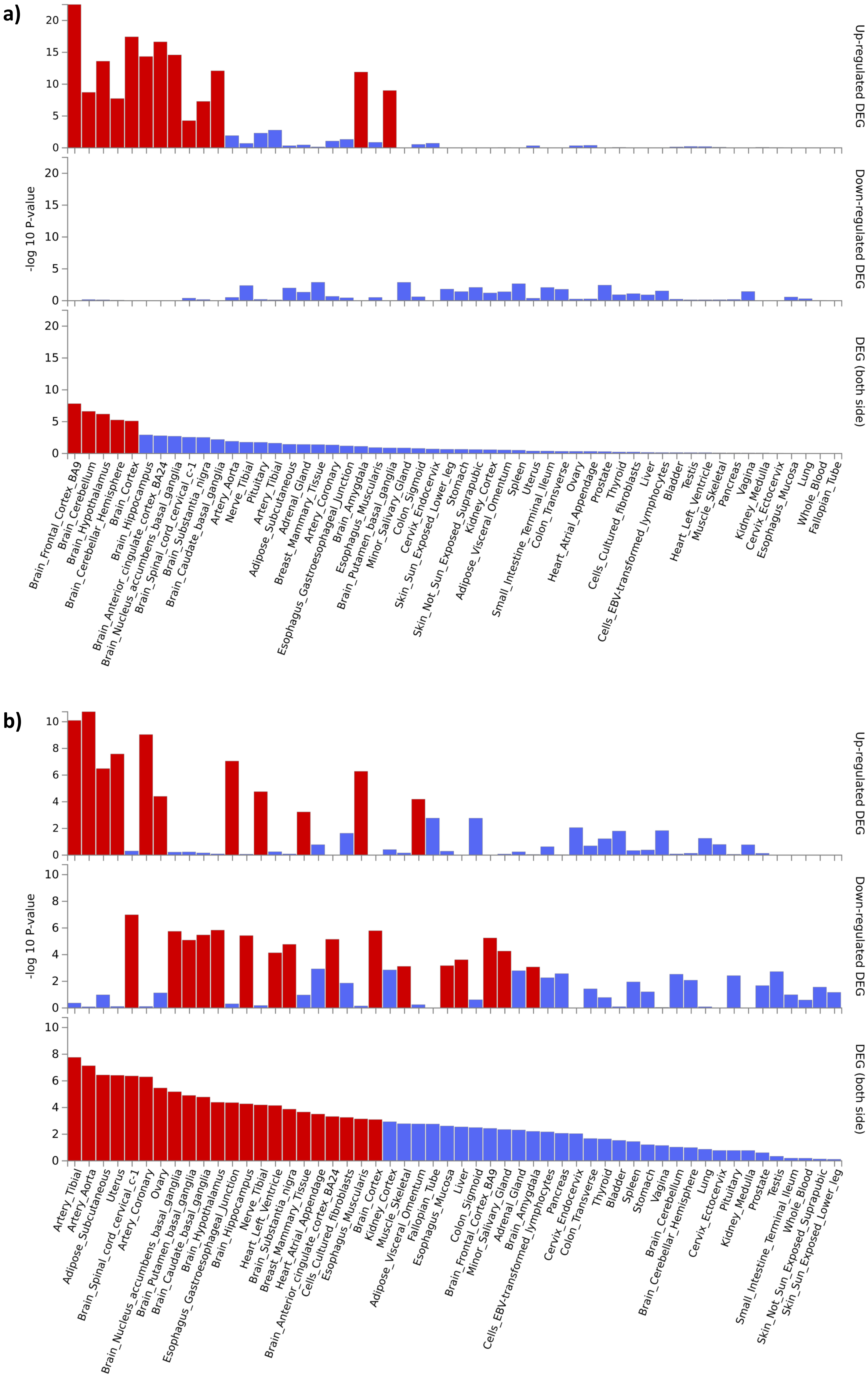


**S Figure 9) Tissue expression profile of genes lying in close proximity of index variants in loci associated with a) overall adiposity or b) body fat distribution traits**

Genes were prioritized for analysis based on lying within 10kb of index variants of loci that were associated with either the overall adiposity or body fat distribution traits. Differentially expressed genes (DEG) were assessed using data from GTEx v8. Significantly enriched DEG sets ($P_{bon} < 0.05$) are highlighted in red.

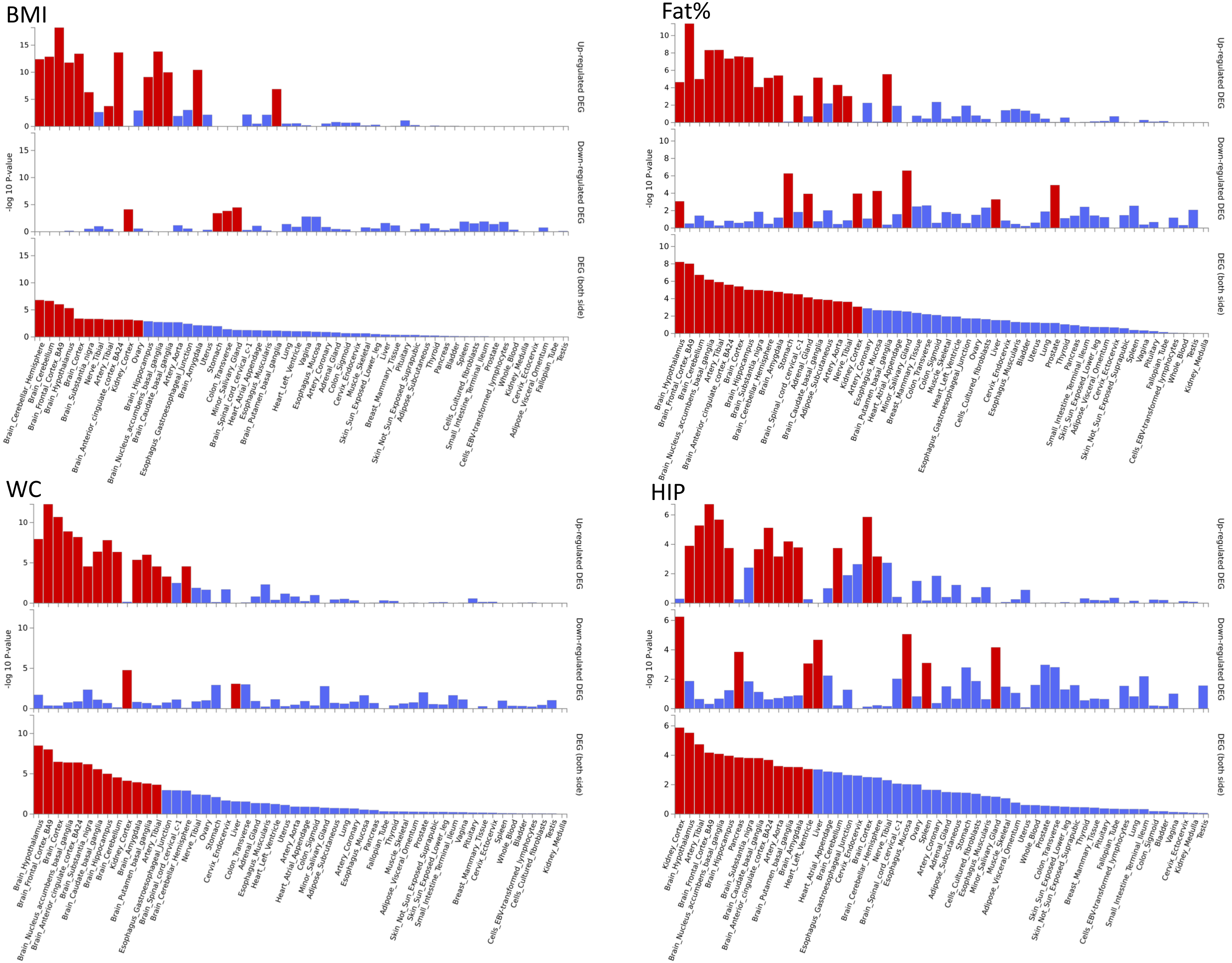


**S Figure 10) Tissue expression profile of genes lying in close proximity of index variants in loci associated with traits classed as overall adiposity traits**

For each trait, genes were prioritized for analysis based on lying within 10kb of index variants of loci that were associated with that trait. Differentially expressed genes (DEG) were assessed using data from GTEx v8. Significantly enriched DEG sets ($P_{bon} < 0.05$) are highlighted in red.

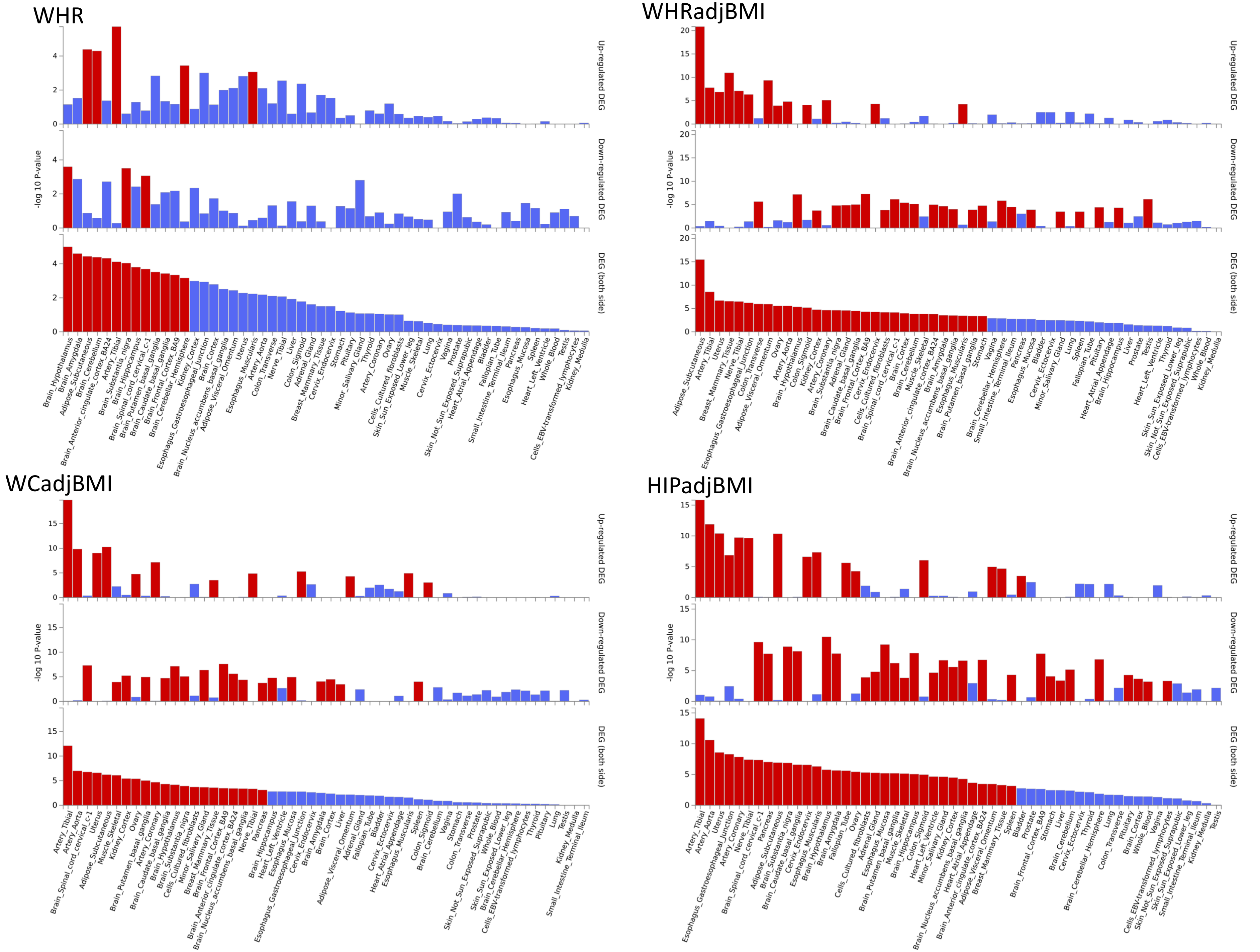


**S Figure 11) Tissue expression profile of genes lying in close proximity of index variants in loci associated with traits classed as body fat distribution traits**

For each trait, genes were prioritized for analysis based on lying within 10kb of index variants of loci that were associated with that trait. Differentially expressed genes (DEG) were assessed using data from GTEx v8. Significantly enriched DEG sets ($P_{bon} < 0.05$) are highlighted in red.

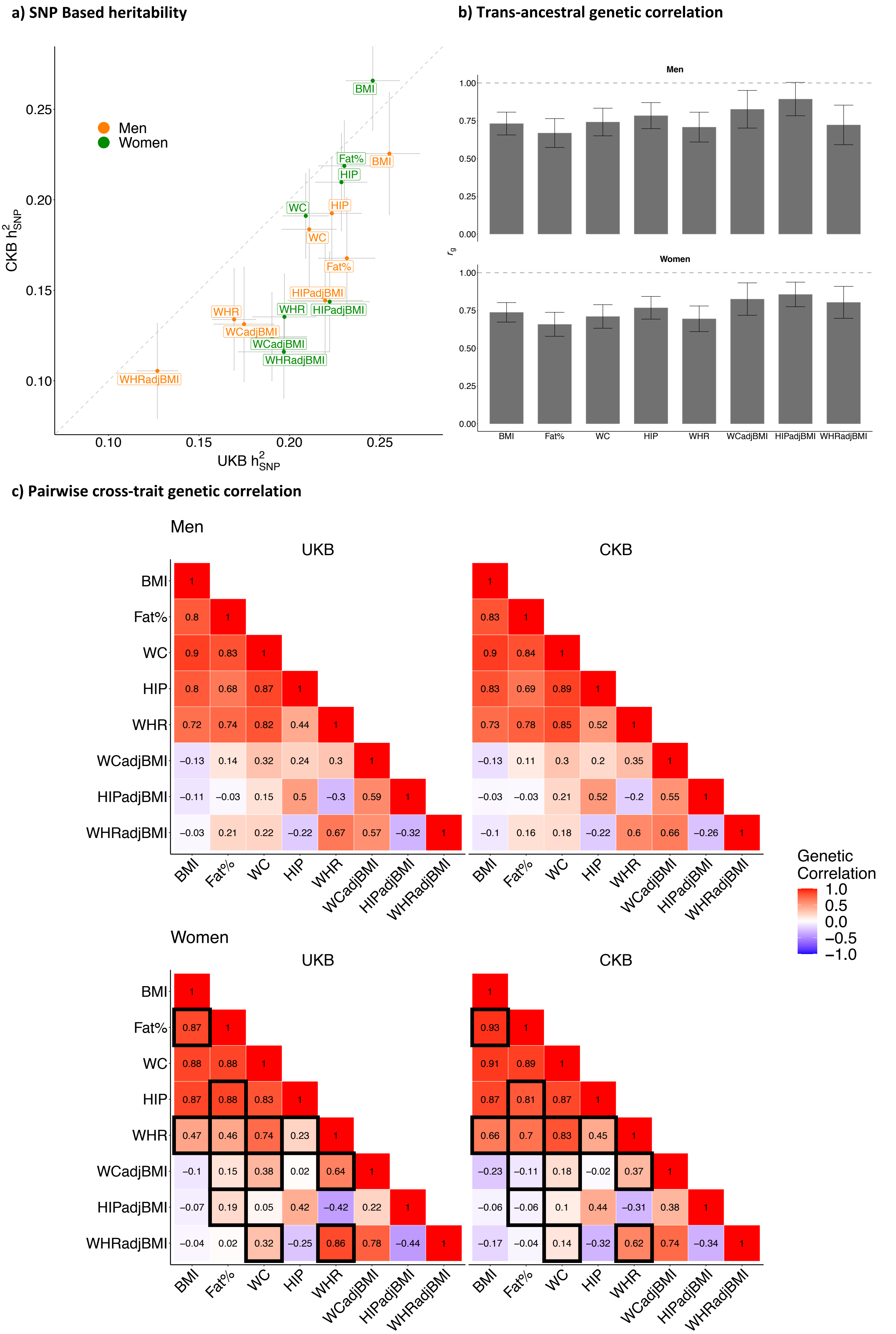


**S Figure 12) Assessment of the sex-specific genetic architecture of the adiposity traits in UKB and CKB**

a) Sex-specific SNP-based heritability estimated from the UKB and CKB sex-specific GWAS results using LDSC regression.
b) Trans-ancestral genetic correlation estimated from the UKB and CKB sex-specific GWAS results using Popcorn.
c) Sex-specific genetic correlation between the adiposity traits estimated from the UKB and CKB sex-specific GWAS results using LDSC regression. Black boxes indicate significant heterogeneity (all P-values < $10^{-3}$) between the biobanks for each sex. The alpha threshold was Bonferroni corrected by the 28 cross-trait comparisons made for the 2 sexes.

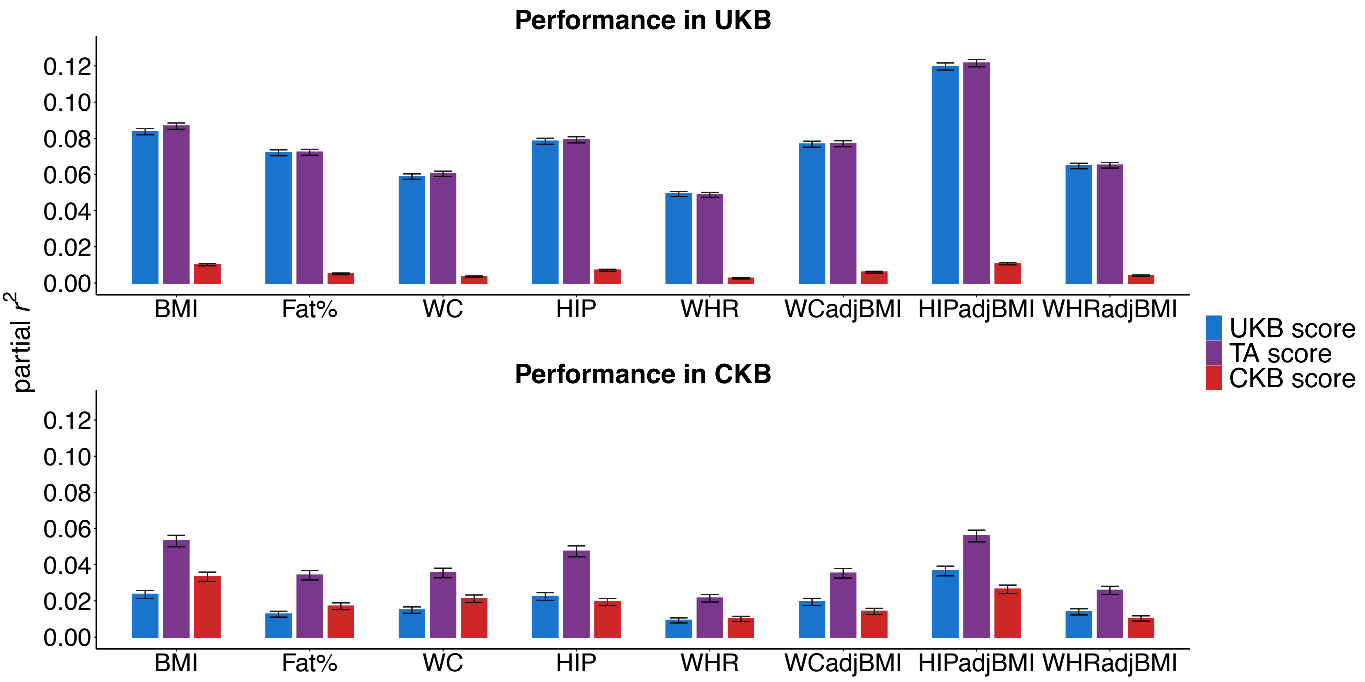


**S Figure 13) Score performance in UKB and CKB for all traits assessed**
Partial $r^2$ values were estimated in the unrelated subset of UKB and CKB from the regression of each adiposity trait against the corresponding score with adjustment for study specific PCs, age, age$^2$ and sex. The 95% CI are plotted based on the standard error of the partial $r^2$ values estimated using Olkin and Finn's approximation.

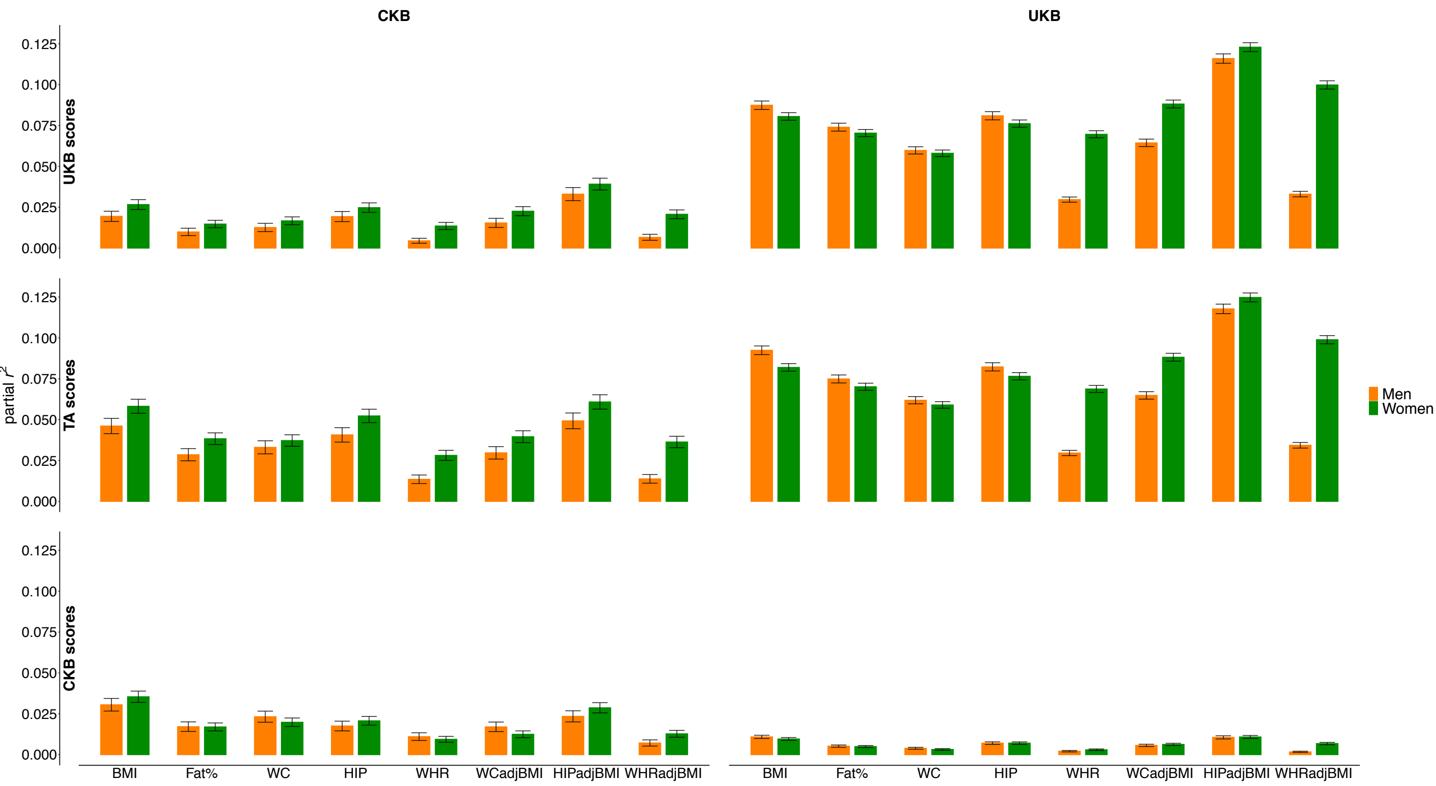


**S Figure 14) Score performance in UKB and CKB stratified by sex**
Partial $r^2$ values were estimated in the unrelated subset of UKB and CKB from the regression of each adiposity trait against the corresponding score with adjustment for study specific PCs, age, age$^2$. The regression was stratified by sex. The 95% CI are plotted based on the standard error of the partial $r^2$ values estimated using Olkin and Finn's approximation.

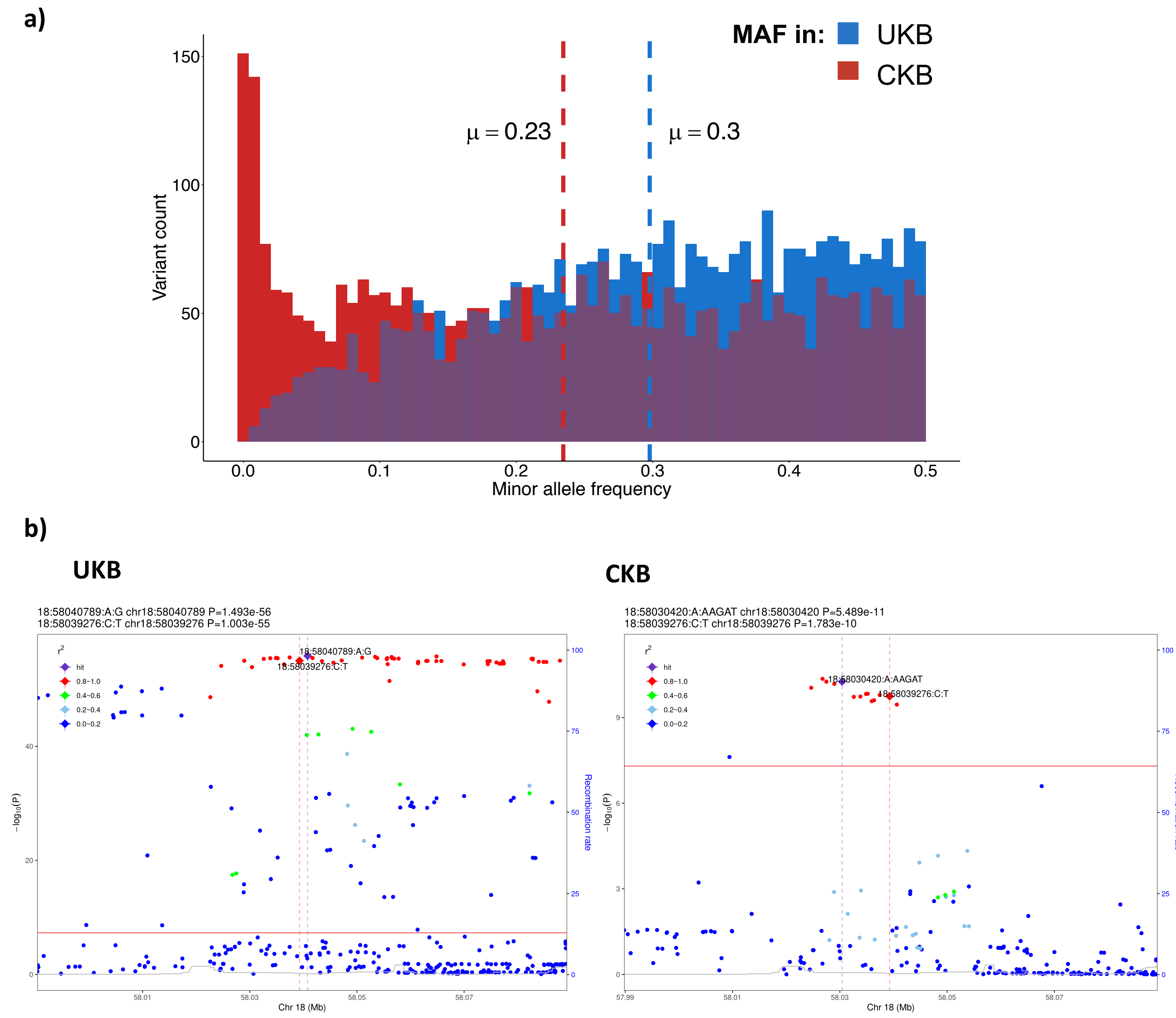


**S Figure 15) MAF and LD differences between UKB and CKB**

a) MAF in UKB and CKB of conditionally independent variants associated with the adiposity traits in UKB. Mean MAF values in each cohort are reported and represented by the dashed lines.

b) Association results and LD at the *MC4R* locus in UKB and CKB, the causal variant at the locus is 18:58039276:C:T. The lead variant at the locus selected in the UKB GWAS was 18:58040789:A:G, while the lead variant selected in CKB was 18:58030420:A:AAGAT.

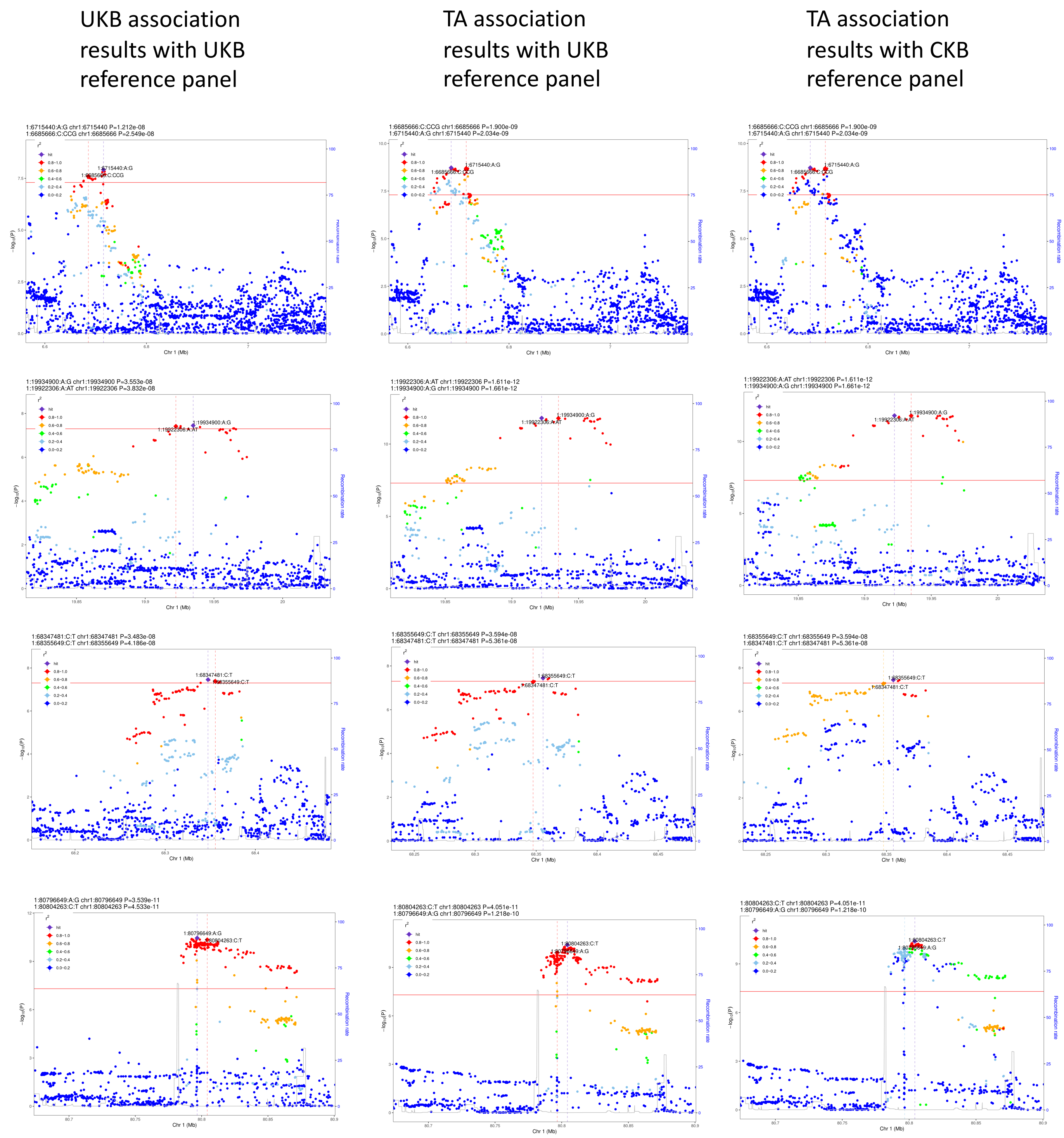


**S Figure 16) Locus zoom plots of regions of the genome where the lead signal changes between the UKB and TA analyses.**

Each row represents a genomic region, the first column is plotted using the UKB association statistics and UKB LD reference panel, the second column is plotted using the TA association statistics and UKB LD reference panel and the third column is plotted using the TA association statistics and CKB LD reference panel. The lead signal in each analysis is colored in purple and the variant detected as the lead signal in the opposing analysis is labelled with it's SNP id.

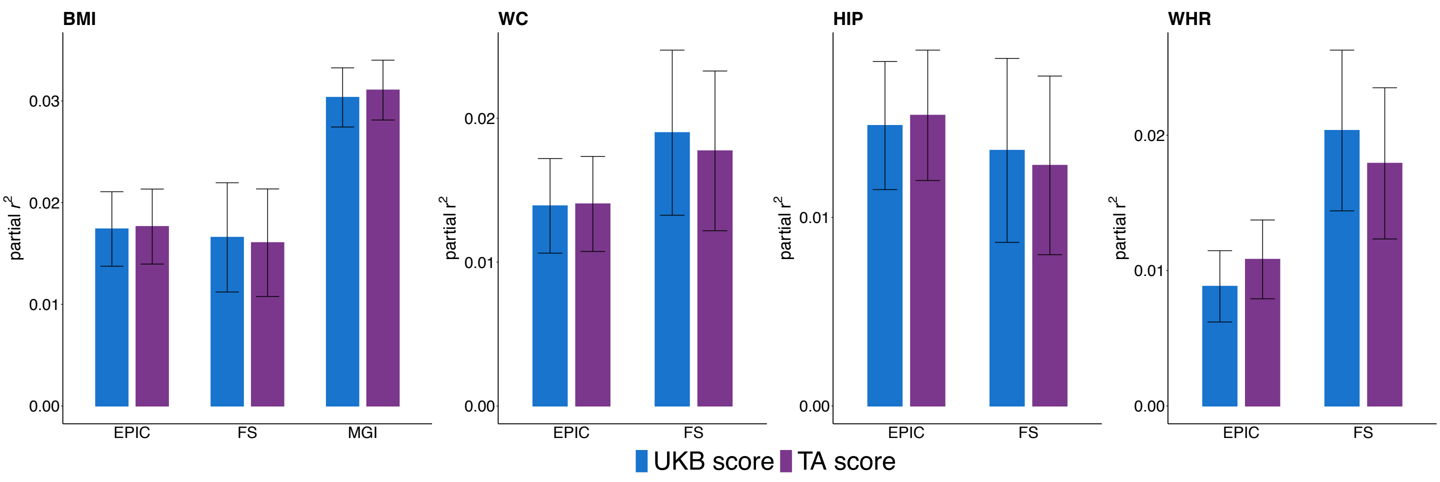


**S Figure 17) Comparison of the predictive performance in EURs of subsets of the UKB and TA scores that are matched by loci.**

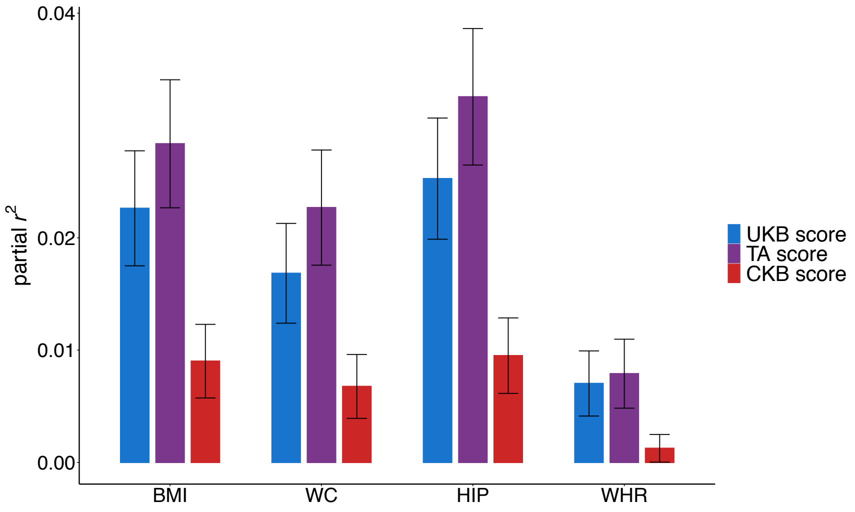


**S Figure 18) Score performance in MCPs participants with low EUR ancestry.**
Individuals in MCPS that have a low proportion ( < 20%) of EUR ancestry were identified using ADMIXTURE analysis. Partial $r^2$ values were estimated in the "low EUR" subset of MCPS from the regression of each adiposity trait against the corresponding score with adjustment for study specific PCs, age, age$^2$ and sex.

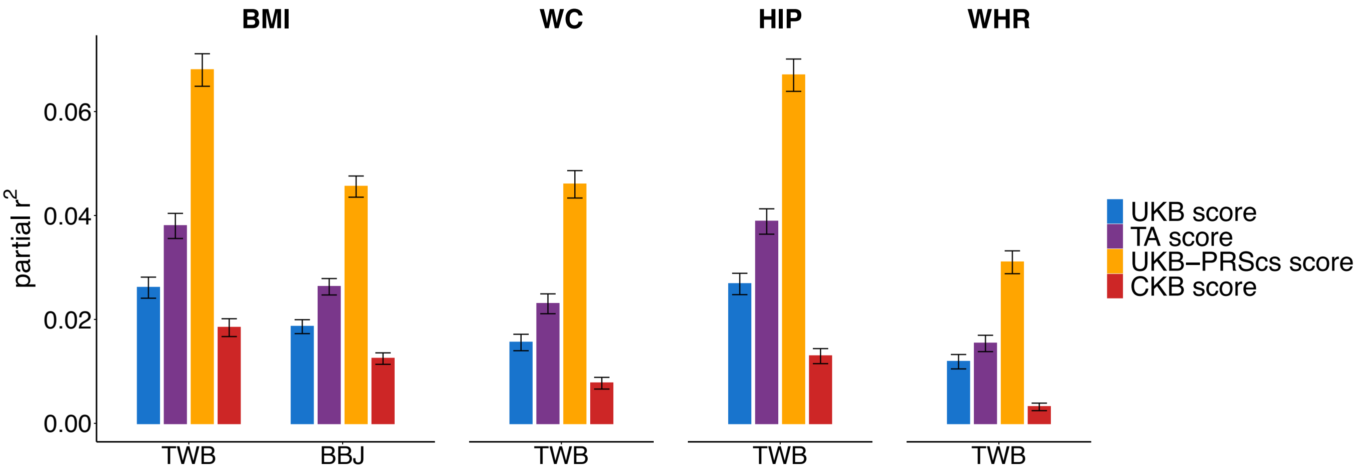


**S Figure 19) Comparison of the UKB, TA and CKB scores against PRScs score performance**
Using the UKB GWAS results and the EUR UKB LD reference dataset provided with the PRScs program we derived a list of variants and weights to be used in the construction of PRScs scores for BMI, WC, HIP and WHR. Partial $r^2$ values were estimated in the unrelated subset of TWB and BBJ from the regression of each adiposity trait against the corresponding score with adjustment for study specific PCs, age, age$^2$ and sex.

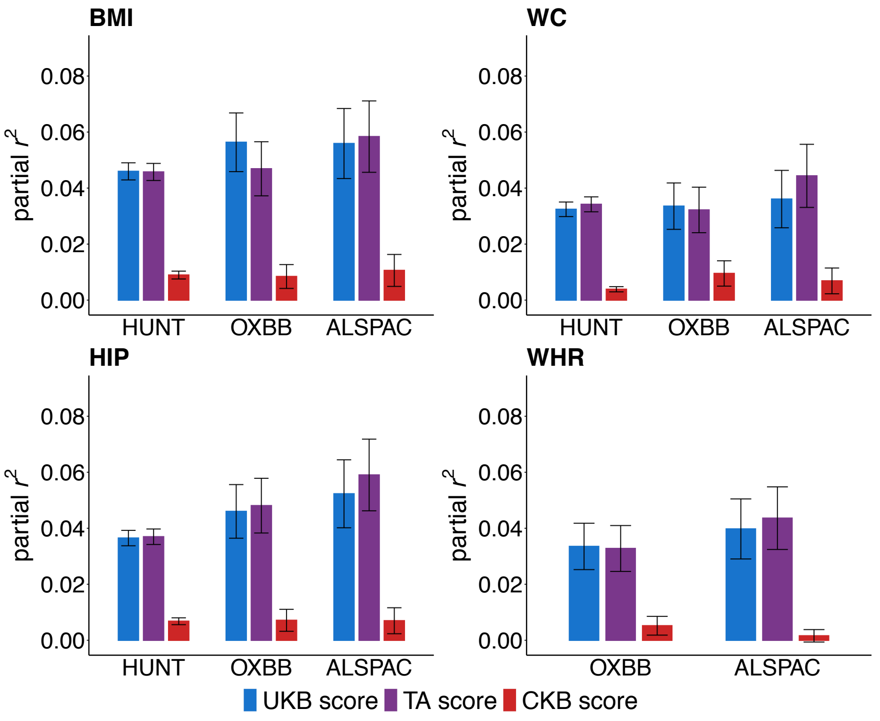


**S Figure 20) Score performance in independent EUR cohorts where 15% or more of score variants were not imputed**
